# Predictive value of circulating NMR metabolic biomarkers for type 2 diabetes risk in the UK Biobank study

**DOI:** 10.1101/2021.10.11.21264833

**Authors:** Fiona Bragg, Eirini Trichia, Diego Aguilar-Ramirez, Jelena Bešević, Sarah Lewington, Jonathan Emberson

## Abstract

**Background:** Effective targeted prevention of type 2 diabetes (T2D) depends on accurate prediction of disease risk. We assessed the role of metabolomic profiling in improving T2D risk prediction beyond conventional risk factors.

**Methods:** NMR-metabolomic profiling was undertaken on baseline plasma samples in 65,684 UK Biobank participants without diabetes and not taking lipid-lowering medication. Cox regression yielded adjusted hazard ratios for the associations of 143 individual metabolic biomarkers (including lipids, lipoproteins, fatty acids, amino acids, ketone bodies and other low molecular weight metabolic biomarkers) and 11 metabolic biomarker principal components (PCs) (accounting for 90% of total variance in individual biomarkers) with incident T2D. These 11 PCs were added to established models for T2D risk prediction, and measures of risk discrimination (c-statistic) and reclassification (continuous net reclassification improvement [NRI], integrated discrimination index [IDI]) were assessed.

**Findings:** During median 11.9 (IQR 11.1-12.6) years’ follow-up, 1719 participants developed T2D. After accounting for multiple testing, 118 metabolic biomarkers showed independent associations with T2D risk (false discovery rate controlled *p*<0.05), of which 103 persisted after additional adjustment for HbA1c. Overall, 10 metabolic biomarker PCs were independently associated with T2D. Addition of PCs to the established risk prediction model (including age, sex, parental history of diabetes, body mass index and HbA1c) improved T2D risk prediction as assessed by the c-statistic (increased from 0.802 [95% CI 0.791-0.812] to 0.830 [0.822-0.841]), continuous NRI (0.44 [0.38-0.49]), and relative (15.0% [10.5%-20.4%]) and absolute (1.5 [1.0-1.9]) IDI.

**Interpretation:** When added to conventional risk factors, circulating NMR-based metabolic biomarkers enhanced T2D risk prediction.

**Funding:** BHF, MRC, CRUK

## Introduction

Both population-level and individual high-risk prevention approaches are essential for addressing the major and rising global public health challenge of type 2 diabetes (T2D). Fundamental to the latter is the ability to accurately predict future T2D risk, enabling targeted or *precision* prevention of the disease,^1^ and ultimately of its complications.^2^ Existing risk prediction models are imperfect, frequently over-estimating T2D risk^3^ and often lacking sufficient specificity to be of use clinically.^4^ Moreover, they characteristically rely on distal risk factors, and consider, at best, only limited molecular pathways. This contrasts with the classical T2D prodrome, comprising dysregulation of multiple molecular pathways over a period of many years.^5^

Through metabolomic profiling, large numbers of biomarkers across multiple biological pathways—proximal and distal—can be quantified in a single measurement, capturing the consequences of genetic variation, environmental influences, and their interactions. Prospective studies have established associations of diverse circulating metabolic biomarkers (e.g., amino acids, fatty acids, hexoses, lipids) with T2D.^6^ As well as providing aetiological insights, these data might feasibly contribute valuable risk prediction information. Previous studies investigating the ability of metabolomics to improve T2D risk prediction over established risk factors have, with the exception of a small number of studies,^7,8^ based their findings on limited T2D cases,^9–14^ frequently investigating only small numbers or single subclasses of metabolic biomarkers,^8–13^ or have used untargeted metabolomic profiling, including unknown biomarkers^13,14^ with limited translational potential. The resulting inconsistent findings leave on-going uncertainty regarding the value of metabolomic profiling for T2D risk prediction.

Using recently-available data from the UK Biobank study, we characterise the prospective associations of circulating metabolic biomarkers, quantified using a high-throughput targeted NMR metabolomics platform, with risk of incident T2D, and examine whether addition of these biomarkers to established models improves prediction of T2D risk.

## Methods

### Study population

Details of the UK Biobank (UKB) study design and population have been described previously.^15^ Briefly, postal invitations to participate were sent to 9.2 million adults aged 40-69 years, living in England, Wales or Scotland and registered with the UK National Health Service. A response rate of 5.5% was achieved, and 502,493 participants were enrolled.

### Data collection

The baseline survey took place between 2006 and 2010 in 22 assessment centres. Self-administered touchscreen questionnaires collected information on sociodemographic and lifestyle factors (including diet, physical activity, smoking and alcohol drinking), and personal (supplemented by verbal interview) and family medical history. Physical measurements, including blood pressure, height, weight, waist circumference (WC) and hip circumference, were undertaken using calibrated instruments with standard protocols. A non-fasting venous blood sample was collected, with the time since last food or drink recorded. After minimal processing at assessment centres, samples were shipped to a central facility for processing and long-term storage at −80°C. Biochemical biomarkers were measured on stored baseline samples at a central UK Biobank laboratory between 2014 and 2017.^16^ These included HDL-cholesterol and triglycerides (AU5400; Beckman Coulter) and HbA1c (VARIANT II TURBO Hemoglobin Testing System; Bio-Rad). Repeat surveys collected the same information as at baseline in addition to certain enhancements; they comprised a resurvey of ~20,000 participants in 2012-13, and an on-going survey of ~100,000 participants which commenced in 2014.^17,18^

All participants consented to be followed-up through linkage to health-related records. These included prior and prospective data on dates and causes of hospital admissions (Hospital Episode Statistics in England, Patient Episode Database for Wales, and Scottish Morbidity Record), and primary care clinical events and prescribing (available for ~45% of participants), as well as date and cause of death obtained from national death registries.

Ethics approval for the UK Biobank was obtained from the North West Multi-centre Research Ethics Committee (Ref: 11/NW/0382). All participants provided informed written consent.

### Metabolic biomarker quantification

A high-throughput NMR-metabolomics platform^19,20^ was used to undertake metabolomic profiling in baseline plasma samples from a randomly-selected subset of ~120,000 UKB participants.^21^ This simultaneously quantified 249 metabolic biomarkers (168 directly-measured and 81 ratios of these), including lipids, fatty acids, amino acids, ketone bodies and other low molecular weight metabolic biomarkers (e.g., gluconeogenesis related metabolites), as well as lipoprotein subclass distribution, particle size and composition. A subset of 143 (**Webtable 1**) were selected for inclusion in the presented analyses, focussing on those which were directly measured and could not be inferred from other biomarkers.

### Assessment of incident type 2 diabetes status

Incident T2D status was ascertained through: i) self-report of T2D diagnosis or glucose-lowering medication use at repeat surveys; ii) coded T2D diagnoses recorded in primary care, hospital admission or death registry data; or iii) glucose-lowering medication prescribing in primary care data (**Webtable 2**). Only those participants without diagnostic codes for other specified diabetes types (type 1/ malnutrition-related/ other specified diabetes) were considered to have T2D.

### Statistical analysis

Analyses excluded those with previously-diagnosed diabetes of any type (based on self-report, primary care or inpatient hospital data), taking regular glucose-lowering medication (based on self-report or primary care data) or with HbA1c ≥6.5% (corresponding to 48 mmol/mol and consistent with undiagnosed diabetes) at the baseline survey. Those with missing or extreme NMR-biomarker or covariate data (see below), or who were taking lipid-lowering medications at recruitment, were also excluded from the main analyses (**Webfigure 1**).

All NMR-biomarkers were log transformed and standardised. Principal component analysis was then employed to reduce the large number of correlated NMR-biomarkers (**Webfigure 2**) to a much smaller number of uncorrelated principal components (PCs) which retained most (>90%) of the variance in the individual biomarkers. Cox regression was used to assess the individual relevance of each NMR-biomarker (and each PC) to risk of incident T2D. First, to examine the shape of the associations, participants were grouped into baseline categories defined by quartiles of their distributions. Subsequently, continuous analyses of each NMR-biomarker (and each PC) were done to estimate the HR per 1-SD higher baseline level. Cox models were stratified by age-at-risk (5-year age groups) and sex, and adjusted for assessment centre (22 centres), Townsend deprivation index (numeric), smoking (4 categories), alcohol drinking (4 categories), body mass index (BMI) (numeric), waist-to-hip ratio (WHR) (numeric), fasting time (numeric) and spectrometer (6 spectrometers). Participants who did not develop incident T2D were censored at the earliest of death, loss to follow-up or 31 December 2020. For significance testing, the Benjamini-Hochberg method was used to control the false discovery rate (FDR).^22^ Sensitivity analyses examined associations separately by age (<55 vs ≥55 years) and sex, and after additional adjustment for other factors (HbA1c, ethnicity, parental history of diabetes, physical activity and dietary factors [whole and refined grains, fruit, vegetables, cheese, unprocessed red meat, processed meat, non-oily and oily fish, type of spread, caffeinated and decaffeinated coffee, tea and dietary supplements]). In addition, the impact of excluding the first three years of follow-up was assessed, and, for the analysis of each PC, mutual adjustment for all preceding PCs.

Then, to assess whether circulating NMR-biomarkers could improve prediction of T2D risk, the selected PCs were added to ‘traditional’ T2D risk prediction models.^23^ Two such models were assessed: a ‘basic’ model, including age (<50, 50-64, ≥65 years), sex, parental history of diabetes, BMI (<25.0, 25.0-29.9, ≥30.0 kg/m^2^) and HbA1c (<6.0% vs ≥6.0%); and an ‘extended’ model, which additionally included blood pressure (≤130/85 mmHg and not taking anti-hypertensive medication vs >130/85 mmHg or taking anti-hypertensive medication), HDL-cholesterol (<1.0 vs ≥1.0 mmol/L in men; <1.3 vs ≥1.3 mmol/L in women), triglycerides (<1.7 vs ≥1.7 mmol/L), and WC (≤102 vs >102 cm in men; ≤88 vs >88 cm in women).^23^ The discriminatory ability of each model before and after including the PCs was assessed using Harrell’s c-statistic,^24^ and the likelihood ratio test was used to compare the fits of nested models (i.e., those including versus excluding the PCs). Relative and absolute integrated discrimination improvement (IDI)^25^ and continuous net reclassification improvement (NRI)^26^ were estimated to assess risk reclassification. To avoid model optimism, bootstrapping was used to create bias-corrected estimates and CIs for the c-statistics, IDI and NRI. To test model calibration, observed T2D event rates for absolute predicted risk deciles were plotted against their predicted event rates, and calibration slopes were estimated using a Cox regression analysis of predicted risk on observed risk. Calibration slopes and their confidence intervals were estimated from 10-fold cross-validation (pooled using inverse variance weighting). Subsequent analyses assessed the performance of the four risk prediction models solely among 13,695 participants taking lipid-lowering medications at baseline.

Analyses were conducted using SAS (version 9.4) and R (version 3.6.2).

## Results

Of the original 502,493 UKB participants, a random subset of 118,036 (23%) had NMR-biomarker data (**Webfigure 1, Webtable 3**). Of these, 65,684 (56%) had no prior diabetes, were not taking lipid-lowering medication and had complete NMR-biomarker (and other) data, and were included in subsequent analyses. The mean (SD) age was 55.2 (8.0) years, and 58% (n= 37,849) were women (**Table 1**). During 0.8 million person-years of follow-up (median 11.9 [IQR 11.1-12.6]), 1719 cases of incident T2D were identified. Participants who developed T2D were more likely to be male and, at the time of recruitment, tended to be older and of lower socioeconomic status than those who did not develop T2D. They also had higher levels of adiposity, were more likely to be current regular smokers, but less likely to be current regular alcohol drinkers, and more frequently had a parental history of diabetes.

**Table 1.**
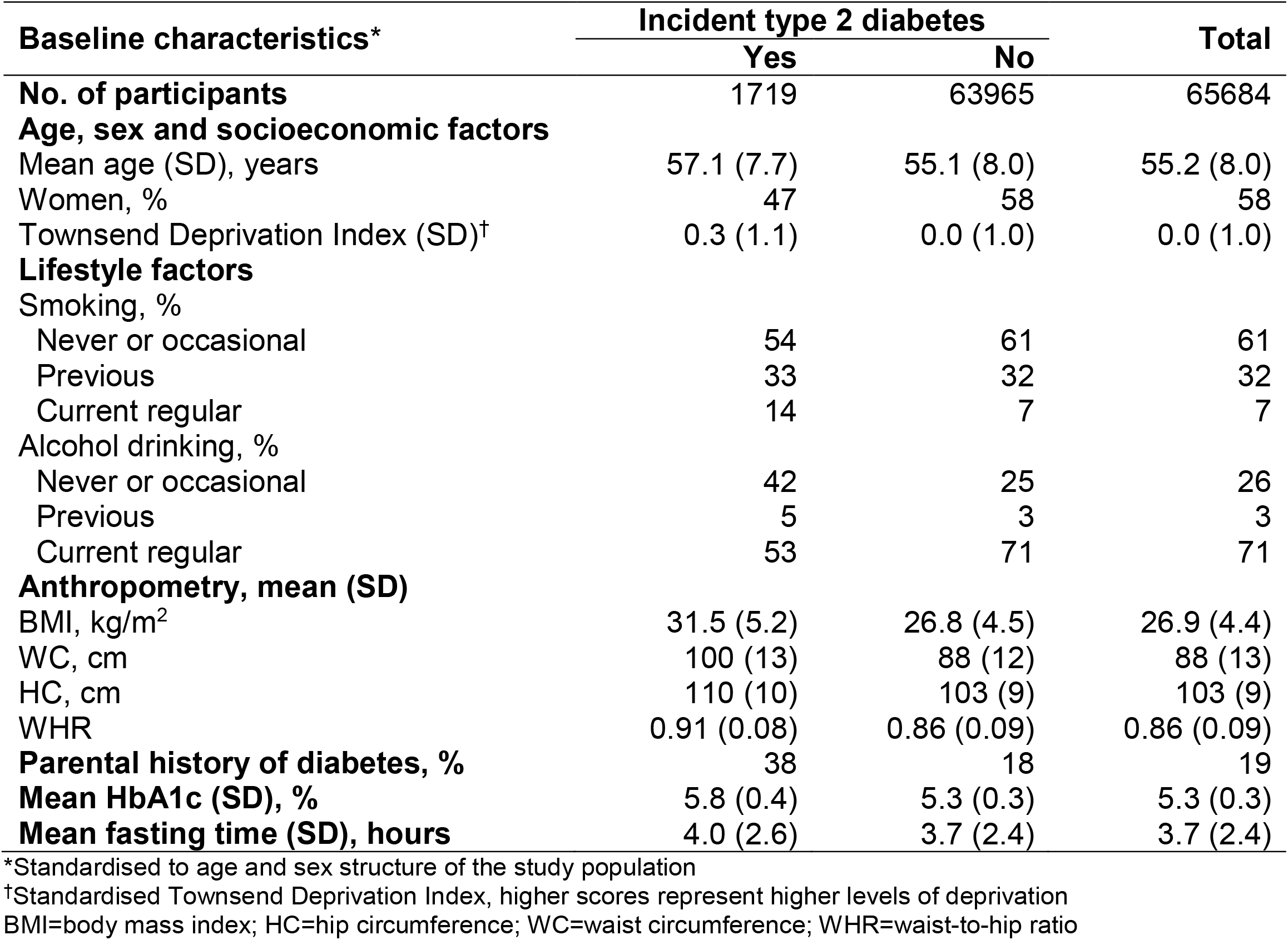
Baseline characteristics by incident type 2 diabetes status.

After adjustment for potential confounding factors and accounting for multiple testing, 118 of the 143 metabolic biomarkers showed statistically significant associations with risk of incident T2D (FDR controlled *p*<0.05) (**Figure 1, Webtable 1, Webfigure 3**). Among the strongest positive associations were those of VLDL particle concentrations, particularly larger VLDL particles, and the lipid concentrations within them. Triglyceride concentrations in all 14 lipoprotein subclasses were also very strongly positively associated with incident T2D. Conversely, concentrations of larger HDL particles, and the cholesterol and phospholipids within those particles, were inversely associated with T2D. Higher branched chain amino acid (BCAA)—leucine, isoleucine and valine—concentrations were associated with higher risk of T2D, as were higher concentrations of alanine, phenylalanine and tyrosine. Glutamine and glycine were inversely associated with T2D. Relative to total fatty acids, higher concentrations of polyunsaturated, omega-3 and omega-6 fatty acids, and of docosahexaenoic and linoleic acids were associated with lower T2D risk, whereas higher concentrations of saturated and monounsaturated fatty acids were associated with higher T2D risk. Higher plasma glycoprotein acetyls, a marker of inflammation, were also associated with higher T2D risk.

**Figure 1.**
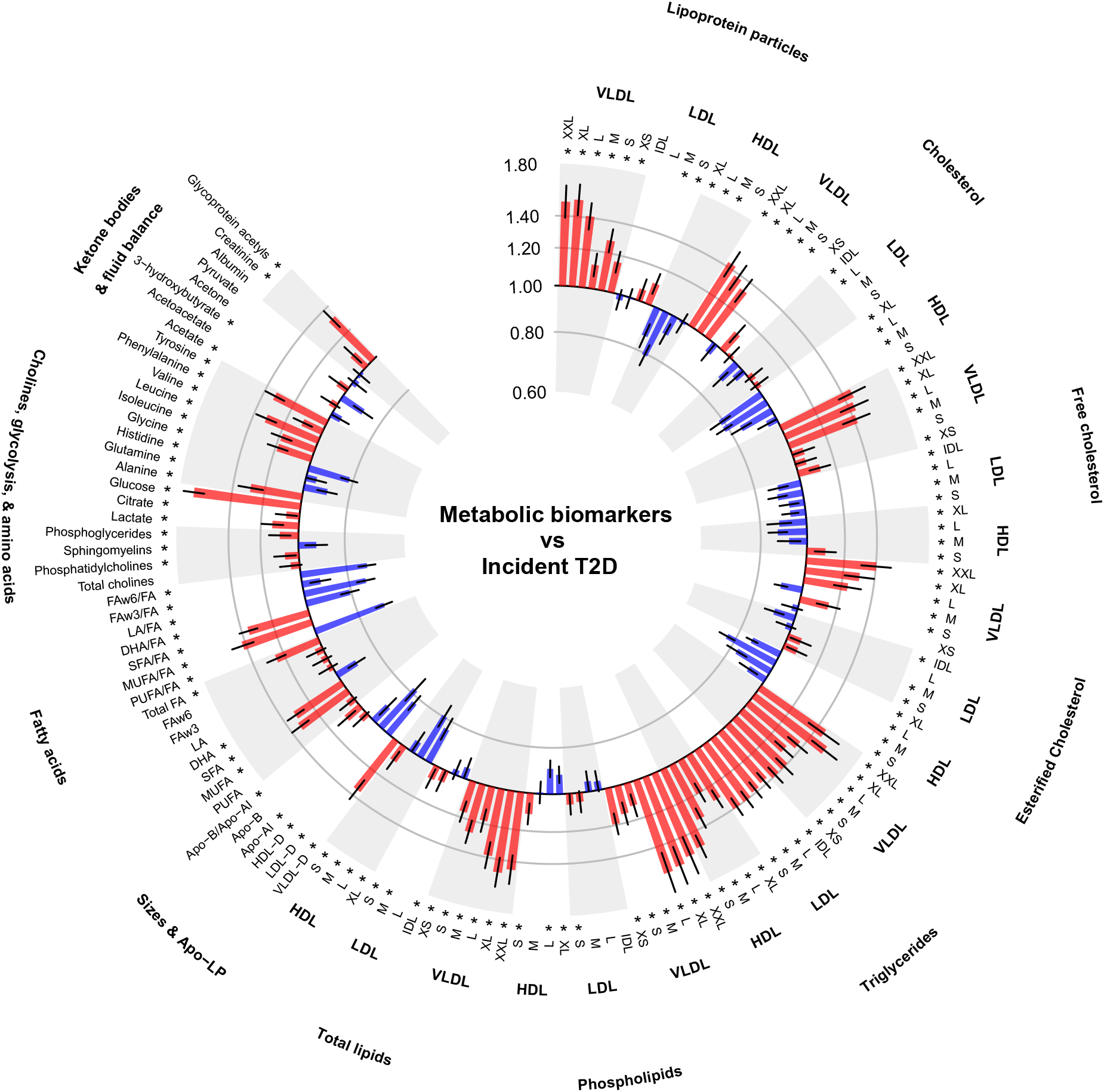
Associations of metabolic biomarkers with risk of incident type 2 diabetes. Hazard ratios (with 95% confidence intervals) are presented per 1−SD higher metabolic biomarker on the natural log scale, stratified by age−at−risk and sex and adjusted for assessment centre, Townsend deprivation index, smoking, drinking, body mass index, waist−to−hip ratio, fasting duration and spectrometer. *false discovery rate controlled p<0.05. Apo−A1=apolipoprotein A1; Apo−B=apolipoprotein B; DHA=docosahexaenoic acid; FA=fatty acids; FAw3=omega−3 fatty acids; FAw6=omega−6 fatty acids; HDL=high density lipoproteins; HDL−D=high density lipoprotein particle diameter; IDL=intermediate density lipoproteins; L=large; LA=linoleic acid; LDL=low density lipoproteins; LDL−D=low density lipoprotein particle diameter; LP=lipoprotein; M=medium; MUFA=monounsaturated fatty acids; PUFA=polyunsaturated fatty acids; S=small; SFA=saturated fatty acids; T2D=type 2 diabetes; VLDL=very low density lipoproteins; VLDL−D=very low density lipoprotein particle diameter; XL=very large; XS=very small; XXL=extremely large.

After additional adjustment for HbA1c, many associations were moderately attenuated but statistically significant associations of most biomarkers (n=103) with T2D remained (**Webtable 1**). Further adjustment for ethnicity, parental history of diabetes, physical activity and dietary factors did not materially alter the associations. There were no marked differences in the relationships between men and women (**Webfigure 4**), by age at baseline (**Webfigure 5**), or after exclusion of the first three years of follow-up (**Webfigure 6**).

The first 11 PCs of the NMR-biomarkers explained 90% of the total variance present in the 143 individual biomarkers (**Webfigure 7**). The PC loadings from these 11 PCs are shown in **Webfigure 8** (the larger a biomarker’s loading, positive or negative, the more it contributes to that PC) and the associations of these PCs with incident T2D are shown in **Webtable 4**. The major contributors to PC1 were the VLDL and LDL particle concentrations and the lipid concentrations within those particles, while for PC2 they included large HDL particles and lipid concentrations within them. PC1 and PC2 showed opposing associations with T2D (adjusted HR 1.23 [95% CI 1.17-1.30] and 0.78 [0.73-0.82], respectively). Biomarkers across multiple molecular pathways, including lipid concentrations in LDL and HDL particles and apolipoprotein concentrations, were prominent contributors to PC3 (HR 1.23 [95% CI 1.18-1.29]). Within PC4 (HR 1.09 [95% CI 1.04-1.15]), loadings were high for small and very large HDL particles and their lipid concentrations, and amino acids were the major contributors to PC5 (1.07 [1.02-1.13]). Fatty acids were dominant in PC6 (0.97 [95% CI 0.92-1.01]) and also PC7 (0.74 [0.70-0.78]), in which ketone bodies also had large factor loadings. Overall, 10 of the 11 PCs were independently associated with incident T2D, and largely remained so after sequential adjustment for preceding PCs (**Webtable 4**).

In the two traditional risk prediction models, all risk factors were strongly and independently associated with T2D risk (**Webtable 5**). Older age, male sex, parental history of diabetes, higher levels of adiposity, blood pressure, HbA1c and triglycerides, and lower HDL-cholesterol concentration were all associated with higher risk. These relationships largely persisted, although with modest attenuation of some, when metabolic biomarker PCs were added. For both models, 8 of the 11 PCs were significantly associated with T2D risk independently of all other risk factors.

The basic T2D risk prediction model (incorporating age, sex, parental history of diabetes, BMI and HbA1c) demonstrated good calibration of observed versus predicted T2D rates across deciles of predicted risk (calibration slope: 0.99 [95% CI 0.95-1.02]) (**Figure 2**). This did not meaningfully change after addition of metabolic biomarker PCs (0.98 [95% CI 0.95-1.02]). **Table 2** summarises measures of model fit and performance. Addition of the PCs to the basic model resulted in a 17% increase in the chi-square statistic, and yielded an increase in the c-statistic from 0.802 (95% CI 0.791-0.812) to 0.830 (0.822-0.841). Improved T2D risk prediction on addition of the PCs was also evidenced by estimates of the overall continuous NRI (0.44 [95% CI 0.38-0.49]), with an improvement of 0.15 (0.12-0.20) in events and 0.28 (0.26-0.31) in non-events, and both absolute (1.5 [1.0-1.9]) and relative (15.0% [10.5%-20.4%]) IDI. The extended model (basic model plus blood pressure, WC, HDL-cholesterol and triglycerides) achieved a c-statistic of 0.829 (95% CI 0.819-0.838). Modest improvements in model fit and performance were observed following addition of metabolic biomarker PCs to this model, with a 6% increase in the chi-square statistic, a c-statistic of 0.837 (95% CI 0.831-0.848), an overall continuous NRI of 0.22 (0.17-0.28), an absolute IDI of 0.7 (0.4-1.1) and a relative IDI of 6.3% (4.1%-9.8%). The extended model was well-calibrated, both with and without inclusion of metabolic biomarker PCs (0.99 [95% CI 0.96-1.02] and 0.98 [0.95-1.01], respectively). When analyses were repeated among participants taking lipid-lowering medications at baseline, c-statistics for the four individual T2D risk prediction models were lower than in the main study population, but estimates of relative performance of the nested models were broadly comparable (**Webtable 6**).

**Table 2.**
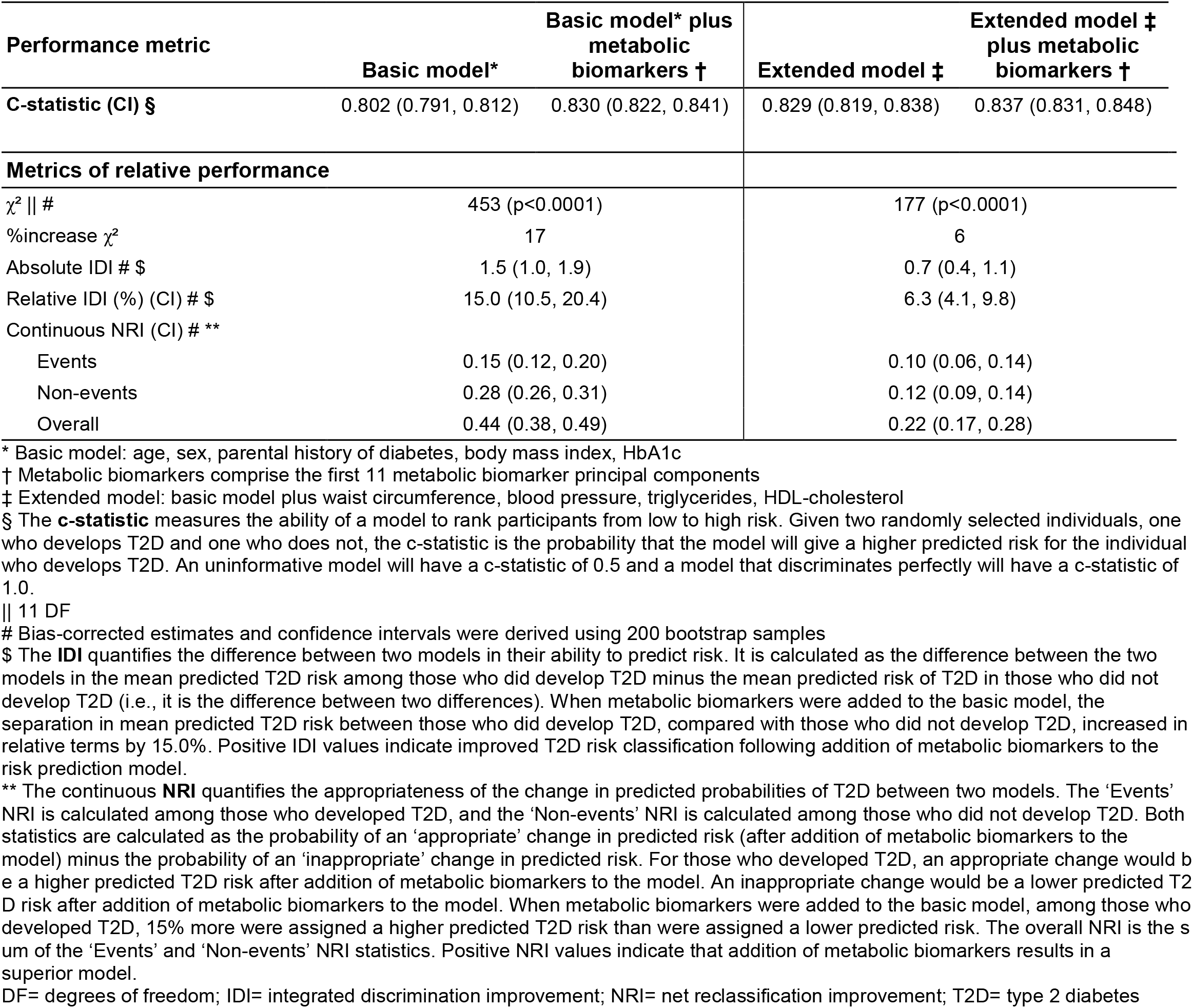
Performance of risk prediction models for incident type 2 diabetes.

**Figure 2.**
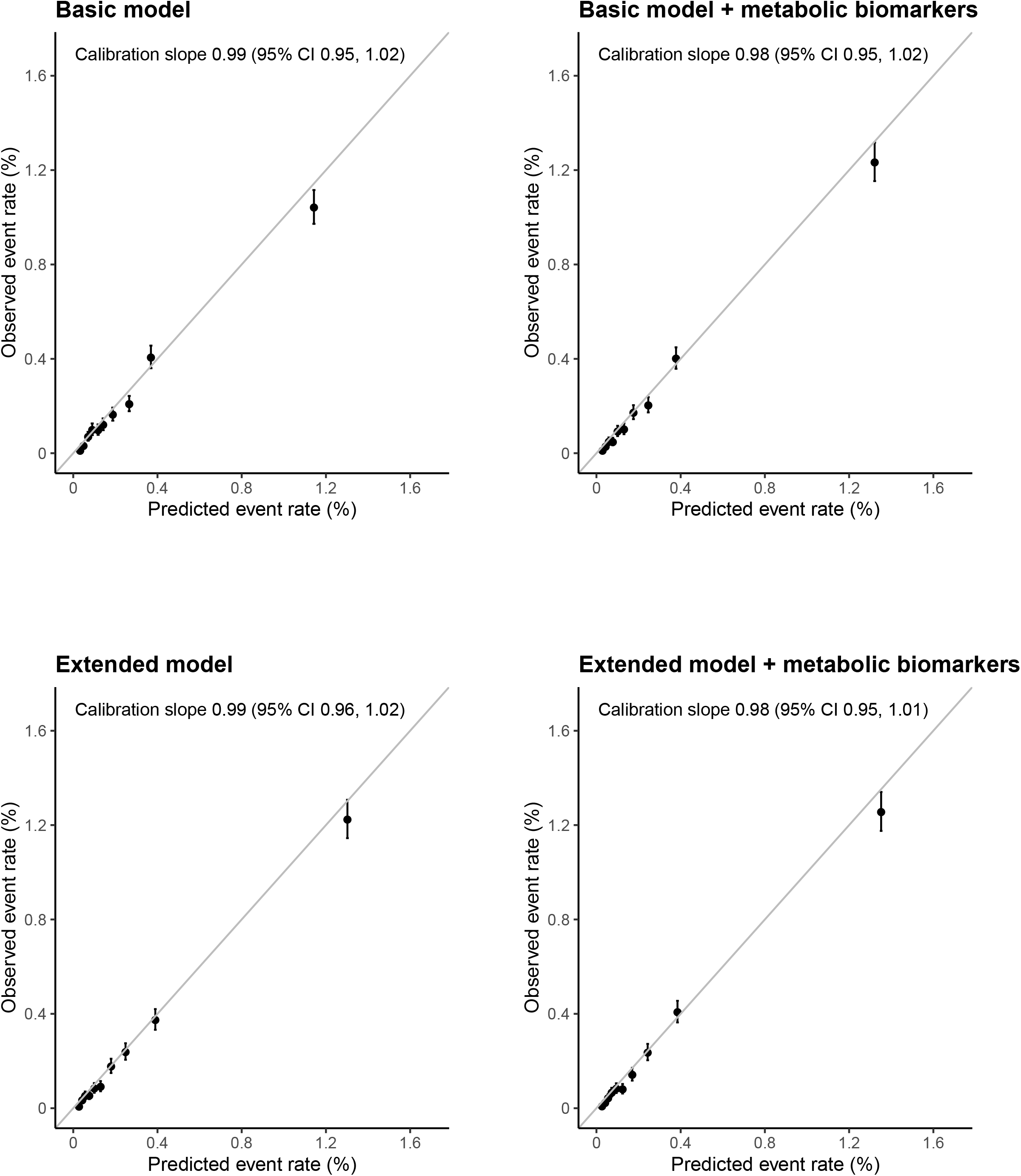
Calibration of prediction models for incident type 2 diabetes from cross−validation. For each model, the observed and predicted T2D event rates are shown for each of 10 equally-sized groups of absolute predicted risk. Vertical lines represent 95% CIs. Calibration slopes are presented from 10−fold cross−validation (pooled using inverse variance weighting) and were derived from a Cox regression of the predicted risk on the observed risk. Basic model: age, sex, parental history of diabetes, body mass index, HbA1c. Extended model: basic model plus waist circumference, triglycerides, and HDL−cholesterol. Metabolic biomarkers comprise the first 11 metabolic biomarker principal components.

## Discussion

This prospective population-based cohort study of over 65,000 middle-aged adults with 1719 cases of new-onset T2D is, to our knowledge, the largest study to-date to examine the predictive value of circulating metabolic biomarkers for T2D risk. Strong independent associations of diverse biomarkers, quantified using targeted NMR-based metabolomic profiling, including lipoprotein particle size and composition, amino acids and fatty acids, with risk of incident T2D were observed. When added to an established risk prediction model comprising basic clinical risk factors and HbA1c, PCs derived from 143 circulating biomarkers substantially improved T2D risk prediction.

Our study found strong positive associations of VLDL particle measures and triglyceride concentrations with incident T2D risk, and inverse associations of HDL particle size and lipids within larger HDL particles. These findings are qualitatively, and broadly quantitatively, consistent with previous studies,^27,28^ and are characteristic of lipoprotein profiles associated with insulin resistance.^29^ This is also thought to underlie the strong positive associations of BCAAs—leucine, isoleucine and valine—with risk of T2D observed in UKB and in previous studies among diverse populations.^6,8,27^ More specifically, genetic association studies have shown increased BCAA levels as a consequence of insulin resistance,^30^ which, in turn, appear to be causally related to T2D.^31^ We replicated findings of studies showing higher levels of phenylalanine, tyrosine and alanine,^6,8,27^ and lower concentrations of glutamate^6,27^ and glycine^6^ several years prior to T2D diagnosis, and the observed T2D-associated fatty acid profiles are broadly consistent with previous investigations.^27,32^ Insulin resistance and inflammation are postulated to underlie some or all of these associations,^6,8,32^ but the nature of the relationships of these and other metabolic biomarkers with T2D, including their causal significance, remains uncertain. Despite this, these findings provide clear evidence of the relevance of diverse metabolic biomarkers to T2D risk.

The basic T2D risk prediction model examined in the present study, which included standard clinical risk predictors and HbA1c, demonstrated good discriminatory ability in the UKB population, yielding a c-statistic (0.80) consistent with that reported for similar models across varied populations.^33^ This highlights one of the major challenges of identifying novel predictive biomarkers for T2D. That is, that established clinical risk factors perform so well in predicting T2D risk that achieving clinically meaningful improvements above and beyond these is difficult. Despite this, addition of metabolic biomarkers to this model improved, albeit modestly, model fit and risk discrimination (c-statistic 0.83). Although some previous studies have observed no improvement in risk discrimination with addition of metabolic biomarkers to similar traditional risk prediction models,^9,12,34^ several have investigated the impact of only limited biomarkers.^9,12^ Inclusion of more diverse biomarkers has tended to achieve greater gains in model discrimination.^7,14,27^ For example, in a case-cohort study in Germany, comprising 800 T2D cases and a randomly-selected subcohort of 2282 adults (mean follow-up 7 years), addition of 14 metabolic biomarkers (including hexoses, amino acids and fatty acids) to an established T2D risk score, comprising clinical risk factors and glycaemia, resulted in moderate, but statistically significant, improvement in risk discrimination (increase in c-statistic from 0.901 to 0.912; p<0.0001).^7^ However, even these studies have tended to investigate highly selected subsets of biomarkers. In contrast, use of principal component analysis in the present study facilitated inclusion of information from all 143 metabolic biomarkers, despite their highly correlated nature. Many individual biomarkers most strongly associated with T2D were prominent contributors to the PCs selected for inclusion in risk prediction models, most of which were associated with incident T2D in fully adjusted regression models.

The only modest, and non-significant, gains in discrimination when metabolic biomarkers were added to the extended T2D risk prediction model in the present study likely reflects overlap between measured metabolic biomarkers and blood-based risk factors included in the extended model, limiting the clinical relevance of this comparison given both assay types would unlikely be used simultaneously. However, it may also reflect the insensitivity of the c-statistic to improvements in predictive performance with addition of new, even strong, risk predictors to established models.^35^ More global measures of model performance provided strong supportive evidence of the value of metabolic biomarkers for T2D risk prediction. Their addition to the basic risk prediction model was associated with improvement in the prediction of T2D using measures of risk reclassification, specifically the IDI and continuous NRI. Of note, the NRI was driven more by reductions in predicted risk among participants who did not develop T2D, suggesting metabolomic profiling may be particularly valuable for reducing unnecessary prevention interventions among individuals at low risk of T2D. Increasing availability of standardised, quantitative, high-throughput metabolomics platforms, such as that used in the current study, underscores the translational potential of these findings. Moreover, the metabolomic profiling data these provide may be of wider clinical relevance (e.g., for diagnosis and risk assessment of other cardiometabolic diseases).^20^

In addition to the large number of incident T2D events, our study has several strengths. An established targeted NMR-metabolomics platform, with existing clinical regulatory approvals,^21^ was used; as well as enabling quantification of diverse biomarkers, this facilitates comparisons between study populations and enhances the potential clinical relevance. Moreover, high levels of correlation between NMR and standard clinical chemistry derived concentrations of a subset of biomarkers (**Webfigure 9**) supports the validity of the approach.^36^ Exclusion of participants taking lipid-lowering medication avoided treatment-associated biases, although the broadly comparable performance of the nested risk prediction models in this subpopulation (with a higher frequency of incident T2D) demonstrates the wider generalisability of our findings. Finally, the cohort study design avoided potential biases and loss of precision which may affect more frequently-used nested case-control and case-cohort designs. However, the study also has limitations. Incident T2D was limited to diagnosed cases; although resulting misclassification would likely underestimate associations of metabolic biomarkers with T2D, the relative improvements in model performance (between models with versus without metabolic biomarkers) should be largely unaffected by misclassification in outcome assessment. Quantification of metabolic biomarkers in non-fasting blood samples may have increased inter- and intra-individual variation in biomarker concentrations, and the lack of repeat measurements prevented assessment of, and adjustment for, the latter. However, the main analyses adjusted for fasting time, which accounts for only a small proportion of variation in plasma metabolic biomarker concentrations,^37^ and biomarker measurements at a single point in time are more relevant in the context of risk prediction. Finally, independent validation of the risk prediction findings was not performed, and, given the lifestyle, and health-related characteristics of the UKB population,^38^ the results may not necessarily be generalisable to other populations at higher risk of future T2D.

In summary, this study provides large-scale evidence of the incremental predictive value of metabolomic profiling for prediction of T2D risk. Addition of data on 143 circulating metabolic biomarkers, with replicated prospective associations with T2D, to an established risk prediction model comprising basic clinical risk factors and HbA1c improved T2D risk discrimination and classification. This serves to illustrate the utility of large-scale biobanks for assessment of the clinical relevance and value of emerging biomarkers. Moreover, given increasing availability, including in clinical settings, of high-throughput, comprehensive, targeted metabolomic profiling, these findings have translational potential for enhanced T2D risk stratification and precision prevention.

## Supporting information

TRIPOD checklist

## Data Availability

The underlying data are open access through application to the UK Biobank, and materials and methods will be made freely available through the UK Biobank as part of this project.

## Acknowledgements

This research used the UK Biobank resource (application number 30418). We thank the participants of UK Biobank for their contribution to the resource. The authors are grateful to Nightingale Health Ltd. for providing early access to UK Biobank NMR metabolomics data during Nightingale Health’s exclusivity period, prior to the resource being made openly available to the scientific community.

## Author contributions

All authors were involved in study design, analysis of data, interpretation, or writing the report.

