## Supplementary material for "Predictive value of circulating NMR metabolic biomarkers for type 2 diabetes risk in the UK Biobank study"

*This page is intentionally left blank*

#### Supplementary Appendix, Table of Contents


| | | |
|---|---|---|
| Webtable 1 | Distribution of metabolic biomarkers and their associations with incident type 2 diabetes | Page 2 |
| Webtable 2 | Diagnosis and medication codes for assessment of type 2 diabetes status in primary and secondary healthcare and death registry records and UK Biobank verbal interview | Page 8 |
| Webtable 3 | Baseline characteristics of UK Biobank participants with and without NMR-metabolomics profiling | Page 10 |
| Webtable 4 | Associations of the first 11 metabolic biomarker principal components with risk of incident type 2 diabetes | Page 11 |
| Webtable 5 | Regression models for risk of incident type 2 diabetes | Page 12 |
| Webtable 6 | Performance of risk prediction models for incident type 2 diabetes among participants taking lipid-lowering medication at recruitment | Page 13 |
| Webfigure 1 | Participant exclusions to derive analysis population | Page 14 |
| Webfigure 2 | Cross-correlations of metabolic biomarkers | Page 15 |
| Webfigure 3 | Associations of metabolic biomarkers with risk of incident type 2 diabetes | Page 16 |
| Webfigure 4 | Associations of metabolic biomarkers with risk of incident type 2 diabetes by sex | Page 23 |
| Webfigure 5 | Associations of metabolic biomarkers with risk of incident type 2 diabetes by age | Page 24 |
| Webfigure 6 | Associations of metabolic biomarkers with risk of incident type 2 diabetes excluding the first three years of follow−up | Page 25 |
| Webfigure 7 | Importance of the first 20 metabolic biomarker principal components | Page 26 |
| Webfigure 8 | Characterisation of the first 11 metabolic biomarker principal components | Page 27 |
| Webfigure 9 | Comparison of biomarkers measured by NMR and routine clinical biochemistry assays | Page 29 |

#### Webtable 1. Distribution of metabolic biomarkers and their associations with incident type 2 diabetes

| Metabolic biomarker | Mean (SD) | Hazard ratio (95% CI) per 1-SD higher level on the natural log scale | | | | | | | |
|---|---|---|---|---|---|---|---|---|---|
| | | Standard model* | | Standard model + HbA1c | | Full model † | | Standard model excluding first 3 years of follow-up | |
| | | HR (95% CI) § | p-value ‖ | HR (95% CI) § | p-value ‖ | HR (95% CI) § | p-value ‖ | HR (95% CI) § | p-value ‖ |
| **Lipoprotein particle concentrations** | | | | | | | | | |
| Chylomicrons and extremely large VLDL particles | $1.7^{-06}$ ($1.5^{-06}$) mmol/L | 1.50 (1.42, 1.58) | $7.2^{-24}$ | 1.35 (1.27, 1.42) | $3.6^{-14}$ | 1.32 (1.22, 1.41) | $2.2^{-08}$ | 1.46 (1.38, 1.54) | $2.9^{-18}$ |
| Very large VLDL particles | $3.5^{-06}$ ($2.3^{-06}$) mmol/L | 1.52 (1.44, 1.59) | $2.6^{-27}$ | 1.34 (1.27, 1.41) | $2.1^{-14}$ | 1.30 (1.21, 1.39) | $4.4^{-08}$ | 1.47 (1.39, 1.55) | $1.5^{-20}$ |
| Large VLDL particles | $1.0^{-05}$ ($5.5^{-06}$) mmol/L | 1.41 (1.35, 1.47) | $4.8^{-25}$ | 1.26 (1.20, 1.33) | $1.8^{-12}$ | 1.24 (1.16, 1.31) | $3.4^{-07}$ | 1.38 (1.31, 1.45) | $4.5^{-19}$ |
| Medium VLDL particles | $3.4^{-05}$ ($1.2^{-05}$) mmol/L | 1.12 (1.07, 1.18) | $7.4^{-05}$ | 1.03 (0.98, 1.09) | $2.7^{-01}$ | 1.03 (0.97, 1.10) | $4.1^{-01}$ | 1.11 (1.06, 1.17) | $5.7^{-04}$ |
| Small VLDL particles | $3.9^{-05}$ ($1.3^{-05}$) mmol/L | 1.27 (1.22, 1.33) | $4.9^{-17}$ | 1.15 (1.10, 1.21) | $9.2^{-07}$ | 1.15 (1.09, 1.22) | $1.0^{-04}$ | 1.26 (1.20, 1.32) | $1.9^{-13}$ |
| Very small VLDL particles | $5.5^{-05}$ ($1.4^{-05}$) mmol/L | 1.16 (1.11, 1.21) | $3.1^{-08}$ | 1.06 (1.01, 1.11) | $3.4^{-02}$ | 1.09 (1.03, 1.16) | $9.4^{-03}$ | 1.16 (1.10, 1.21) | $6.0^{-07}$ |
| IDL particles | $3.1^{-04}$ ($7.6^{-05}$) mmol/L | 0.97 (0.93, 1.02) | $3.3^{-01}$ | 0.92 (0.87, 0.97) | $1.5^{-03}$ | 0.94 (0.88, 1.00) | $8.2^{-02}$ | 0.98 (0.93, 1.03) | $4.6^{-01}$ |
| Large LDL particles | $7.3^{-04}$ ($1.7^{-04}$) mmol/L | 0.99 (0.94, 1.04) | $8.1^{-01}$ | 0.94 (0.89, 0.99) | $1.4^{-02}$ | 0.95 (0.89, 1.01) | $1.2^{-01}$ | 1.00 (0.95, 1.05) | $9.6^{-01}$ |
| Medium LDL particles | $2.9^{-04}$ ($7.5^{-05}$) mmol/L | 1.06 (1.01, 1.11) | $4.2^{-02}$ | 0.98 (0.93, 1.03) | $4.4^{-01}$ | 0.99 (0.93, 1.05) | $7.8^{-01}$ | 1.06 (1.01, 1.12) | $4.3^{-02}$ |
| Small LDL particles | $1.7^{-04}$ ($3.7^{-05}$) mmol/L | 1.11 (1.06, 1.16) | $1.1^{-04}$ | 1.02 (0.97, 1.07) | $5.8^{-01}$ | 1.02 (0.96, 1.09) | $5.4^{-01}$ | 1.11 (1.06, 1.17) | $3.5^{-04}$ |
| Very large HDL particles | $2.3^{-04}$ ($9.5^{-05}$) mmol/L | 0.86 (0.80, 0.92) | $3.8^{-06}$ | 0.87 (0.80, 0.93) | $1.4^{-05}$ | 0.92 (0.85, 1.00) | $6.7^{-02}$ | 0.87 (0.80, 0.93) | $4.5^{-05}$ |
| Large HDL particles | $1.4^{-03}$ ($7.6^{-04}$) mmol/L | 0.79 (0.74, 0.85) | $6.7^{-17}$ | 0.83 (0.78, 0.89) | $2.4^{-10}$ | 0.87 (0.80, 0.94) | $4.1^{-04}$ | 0.79 (0.74, 0.85) | $1.3^{-14}$ |
| Medium HDL particles | $3.8^{-03}$ ($9.2^{-04}$) mmol/L | 0.91 (0.85, 0.96) | $7.5^{-04}$ | 0.94 (0.88, 1.00) | $3.9^{-02}$ | 0.98 (0.91, 1.04) | $5.4^{-01}$ | 0.90 (0.84, 0.95) | $4.3^{-04}$ |
| Small HDL particles | $9.6^{-03}$ ($1.3^{-03}$) mmol/L | 0.99 (0.94, 1.04) | $7.0^{-01}$ | 0.97 (0.92, 1.02) | $3.2^{-01}$ | 0.97 (0.91, 1.04) | $4.5^{-01}$ | 0.98 (0.92, 1.04) | $5.1^{-01}$ |
| **Cholesterol concentrations** | | | | | | | | | |
| Chylomicrons and extremely large VLDL particles | 0.06 (0.04) mmol/L | 1.43 (1.36, 1.50) | $6.2^{-23}$ | 1.30 (1.23, 1.37) | $4.1^{-13}$ | 1.26 (1.18, 1.35) | $2.2^{-07}$ | 1.40 (1.33, 1.48) | $1.0^{-17}$ |
| Very large VLDL particles | 0.05 (0.03) mmol/L | 1.38 (1.31, 1.45) | $2.6^{-18}$ | 1.23 (1.16, 1.30) | $1.8^{-08}$ | 1.20 (1.11, 1.28) | $1.1^{-04}$ | 1.35 (1.27, 1.43) | $4.4^{-14}$ |
| Large VLDL particles | 0.10 (0.05) mmol/L | 1.33 (1.26, 1.40) | $3.0^{-16}$ | 1.20 (1.13, 1.26) | $2.8^{-07}$ | 1.17 (1.09, 1.25) | $3.9^{-04}$ | 1.31 (1.24, 1.38) | $1.6^{-12}$ |
| Medium VLDL particles | 0.17 (0.07) mmol/L | 0.95 (0.90, 1.00) | $3.7^{-02}$ | 0.90 (0.86, 0.95) | $6.9^{-05}$ | 0.92 (0.86, 0.98) | $7.5^{-03}$ | 0.95 (0.90, 1.00) | $7.6^{-02}$ |
| Small VLDL particles | 0.16 (0.05) mmol/L | 1.11 (1.06, 1.17) | $2.4^{-04}$ | 1.02 (0.97, 1.08) | $4.6^{-01}$ | 1.03 (0.96, 1.10) | $4.4^{-01}$ | 1.11 (1.05, 1.17) | $8.3^{-04}$ |
| Very small VLDL particles | 0.18 (0.05) mmol/L | 1.01 (0.96, 1.07) | $5.9^{-01}$ | 0.95 (0.90, 1.01) | $9.8^{-02}$ | 0.99 (0.93, 1.05) | $7.9^{-01}$ | 1.02 (0.96, 1.07) | $5.5^{-01}$ |
| IDL particles | 0.83 (0.21) mmol/L | 0.90 (0.85, 0.95) | $2.4^{-05}$ | 0.87 (0.82, 0.92) | $1.7^{-08}$ | 0.90 (0.84, 0.96) | $7.1^{-04}$ | 0.90 (0.85, 0.96) | $2.9^{-04}$ |
| Large LDL particles | 1.11 (0.28) mmol/L | 0.95 (0.90, 1.00) | $3.5^{-02}$ | 0.90 (0.86, 0.95) | $6.1^{-05}$ | 0.93 (0.87, 0.98) | $1.5^{-02}$ | 0.95 (0.90, 1.01) | $8.6^{-02}$ |
| Medium LDL particles | 0.42 (0.12) mmol/L | 1.03 (0.98, 1.08) | $3.3^{-01}$ | 0.96 (0.91, 1.01) | $1.2^{-01}$ | 0.97 (0.91, 1.03) | $3.8^{-01}$ | 1.03 (0.98, 1.09) | $3.1^{-01}$ |
| Small LDL particles | 0.18 (0.04) mmol/L | 1.02 (0.97, 1.07) | $3.8^{-01}$ | 0.95 (0.90, 1.00) | $8.8^{-02}$ | 0.96 (0.90, 1.02) | $3.0^{-01}$ | 1.03 (0.97, 1.08) | $3.4^{-01}$ |
| Very large HDL particles | 0.08 (0.03) mmol/L | 0.79 (0.73, 0.84) | $4.6^{-16}$ | 0.80 (0.74, 0.86) | $9.3^{-13}$ | 0.84 (0.77, 0.91) | $1.5^{-05}$ | 0.79 (0.73, 0.86) | $1.6^{-12}$ |

#### Webtable 1. Distribution of metabolic biomarkers and their associations with incident type 2 diabetes

| Metabolic biomarker | Mean (SD) | Hazard ratio (95% CI) per 1-SD higher level on the natural log scale | | | | | | | |
|---|---|---|---|---|---|---|---|---|---|
| | | Standard model* | | Standard model + HbA1c | | Full model † | | Standard model excluding first 3 years of follow-up | |
| | | HR (95% CI) § | p-value ‖ | HR (95% CI) § | p-value ‖ | HR (95% CI) § | p-value ‖ | HR (95% CI) § | p-value ‖ |
| Large HDL particles | 0.29 (0.17) mmol/L | 0.78 (0.74, 0.83) | $6.4^{-28}$ | 0.82 (0.77, 0.87) | $2.6^{-16}$ | 0.85 (0.79, 0.91) | $5.5^{-07}$ | 0.79 (0.74, 0.84) | $4.5^{-23}$ |
| Medium HDL particles | 0.49 (0.12) mmol/L | 0.84 (0.79, 0.90) | $2.2^{-09}$ | 0.89 (0.83, 0.94) | $3.6^{-05}$ | 0.92 (0.85, 0.98) | $2.1^{-02}$ | 0.84 (0.78, 0.90) | $7.8^{-09}$ |
| Small HDL particles | 0.44 (0.06) mmol/L | 0.95 (0.90, 1.00) | $6.4^{-02}$ | 0.94 (0.89, 0.99) | $3.2^{-02}$ | 0.94 (0.88, 1.01) | $1.1^{-01}$ | 0.94 (0.89, 1.00) | $5.7^{-02}$ |
| **Free cholesterol concentrations** | | | | | | | | | |
| Chylomicrons and extremely large VLDL particles | 0.03 (0.02) mmol/L | 1.46 (1.39, 1.53) | $1.3^{-25}$ | 1.33 (1.26, 1.40) | $2.5^{-15}$ | 1.30 (1.21, 1.38) | $6.1^{-09}$ | 1.43 (1.35, 1.50) | $1.1^{-19}$ |
| Very large VLDL particles | 0.02 (0.01) mmol/L | 1.44 (1.37, 1.51) | $7.3^{-23}$ | 1.28 (1.21, 1.35) | $1.2^{-11}$ | 1.24 (1.16, 1.33) | $1.8^{-06}$ | 1.40 (1.33, 1.48) | $2.6^{-17}$ |
| Large VLDL particles | 0.05 (0.02) mmol/L | 1.42 (1.35, 1.49) | $2.0^{-22}$ | 1.26 (1.19, 1.33) | $7.3^{-11}$ | 1.23 (1.14, 1.31) | $5.0^{-06}$ | 1.39 (1.31, 1.46) | $3.8^{-17}$ |
| Medium VLDL particles | 0.08 (0.03) mmol/L | 1.06 (1.00, 1.11) | $4.6^{-02}$ | 0.99 (0.93, 1.04) | $6.1^{-01}$ | 0.99 (0.93, 1.06) | $8.3^{-01}$ | 1.06 (1.00, 1.11) | $8.2^{-02}$ |
| Small VLDL particles | 0.06 (0.02) mmol/L | 1.05 (1.00, 1.10) | $6.9^{-02}$ | 0.98 (0.92, 1.03) | $4.2^{-01}$ | 0.99 (0.92, 1.05) | $7.7^{-01}$ | 1.05 (1.00, 1.11) | $9.5^{-02}$ |
| Very small VLDL particles | 0.06 (0.01) mmol/L | 1.11 (1.06, 1.17) | $7.4^{-05}$ | 1.03 (0.98, 1.08) | $3.5^{-01}$ | 1.06 (0.99, 1.12) | $1.1^{-01}$ | 1.11 (1.06, 1.17) | $3.4^{-04}$ |
| IDL particles | 0.22 (0.06) mmol/L | 0.89 (0.85, 0.94) | $9.1^{-06}$ | 0.86 (0.81, 0.91) | $1.1^{-09}$ | 0.89 (0.83, 0.95) | $1.5^{-04}$ | 0.90 (0.85, 0.96) | $2.7^{-04}$ |
| Large LDL particles | 0.29 (0.08) mmol/L | 0.88 (0.84, 0.93) | $3.5^{-07}$ | 0.86 (0.82, 0.91) | $3.3^{-10}$ | 0.88 (0.83, 0.94) | $4.2^{-05}$ | 0.89 (0.84, 0.94) | $2.0^{-05}$ |
| Medium LDL particles | 0.12 (0.03) mmol/L | 0.93 (0.88, 0.97) | $2.3^{-03}$ | 0.89 (0.84, 0.93) | $9.6^{-07}$ | 0.90 (0.85, 0.96) | $9.8^{-04}$ | 0.94 (0.89, 0.99) | $1.6^{-02}$ |
| Small LDL particles | 0.05 (0.01) mmol/L | 0.93 (0.89, 0.97) | $3.6^{-04}$ | 0.89 (0.85, 0.93) | $7.7^{-08}$ | 0.91 (0.86, 0.96) | $4.8^{-04}$ | 0.94 (0.89, 0.98) | $5.0^{-03}$ |
| Very large HDL particles | 0.02 (0.01) mmol/L | 0.88 (0.82, 0.93) | $1.6^{-05}$ | 0.87 (0.81, 0.93) | $1.3^{-05}$ | 0.91 (0.84, 0.98) | $2.2^{-02}$ | 0.89 (0.82, 0.95) | $4.0^{-04}$ |
| Large HDL particles | 0.07 (0.04) mmol/L | 0.87 (0.82, 0.91) | $2.5^{-09}$ | 0.89 (0.84, 0.93) | $2.3^{-06}$ | 0.92 (0.86, 0.99) | $3.4^{-02}$ | 0.87 (0.82, 0.92) | $4.0^{-08}$ |
| Medium HDL particles | 0.09 (0.02) mmol/L | 0.92 (0.86, 0.97) | $2.9^{-03}$ | 0.94 (0.88, 0.99) | $3.1^{-02}$ | 0.98 (0.91, 1.05) | $6.1^{-01}$ | 0.91 (0.85, 0.97) | $1.9^{-03}$ |
| Small HDL particles | 0.11 (0.02) mmol/L | 1.09 (1.04, 1.14) | $8.9^{-04}$ | 1.05 (0.99, 1.10) | $1.1^{-01}$ | 1.07 (1.01, 1.13) | $4.7^{-02}$ | 1.08 (1.02, 1.13) | $1.0^{-02}$ |
| **Esterified cholesterol concentrations** | | | | | | | | | |
| Chylomicrons and extremely large VLDL particles | 0.03 (0.02) mmol/L | 1.40 (1.33, 1.47) | $9.0^{-20}$ | 1.27 (1.20, 1.34) | $7.3^{-11}$ | 1.23 (1.15, 1.32) | $5.7^{-06}$ | 1.37 (1.30, 1.45) | $2.5^{-15}$ |
| Very large VLDL particles | 0.03 (0.01) mmol/L | 1.30 (1.23, 1.37) | $1.3^{-12}$ | 1.16 (1.09, 1.23) | $4.2^{-05}$ | 1.13 (1.05, 1.22) | $7.4^{-03}$ | 1.28 (1.20, 1.36) | $6.9^{-10}$ |
| Large VLDL particles | 0.05 (0.02) mmol/L | 1.24 (1.17, 1.30) | $5.9^{-10}$ | 1.13 (1.06, 1.19) | $5.5^{-04}$ | 1.11 (1.03, 1.19) | $2.0^{-02}$ | 1.22 (1.15, 1.29) | $8.7^{-08}$ |
| Medium VLDL particles | 0.09 (0.04) mmol/L | 0.90 (0.87, 0.93) | $1.7^{-10}$ | 0.88 (0.85, 0.91) | $5.5^{-14}$ | 0.90 (0.85, 0.94) | $1.6^{-06}$ | 0.90 (0.87, 0.94) | $1.3^{-08}$ |
| Small VLDL particles | 0.10 (0.03) mmol/L | 1.15 (1.09, 1.21) | $3.4^{-06}$ | 1.05 (1.00, 1.11) | $1.1^{-01}$ | 1.06 (0.99, 1.13) | $1.5^{-01}$ | 1.15 (1.09, 1.21) | $2.5^{-05}$ |
| Very small VLDL particles | 0.13 (0.04) mmol/L | 0.97 (0.92, 1.02) | $3.1^{-01}$ | 0.93 (0.88, 0.98) | $3.6^{-03}$ | 0.96 (0.90, 1.02) | $2.6^{-01}$ | 0.98 (0.92, 1.03) | $4.4^{-01}$ |
| IDL particles | 0.61 (0.16) mmol/L | 0.90 (0.85, 0.95) | $3.9^{-05}$ | 0.87 (0.83, 0.92) | $5.4^{-08}$ | 0.90 (0.85, 0.96) | $1.3^{-03}$ | 0.91 (0.85, 0.96) | $3.4^{-04}$ |
| Large LDL particles | 0.82 (0.21) mmol/L | 0.97 (0.92, 1.02) | $2.8^{-01}$ | 0.92 (0.87, 0.97) | $1.5^{-03}$ | 0.94 (0.88, 1.00) | $6.9^{-02}$ | 0.98 (0.92, 1.03) | $4.2^{-01}$ |

#### Webtable 1. Distribution of metabolic biomarkers and their associations with incident type 2 diabetes

| Metabolic biomarker | Mean (SD) | Hazard ratio (95% CI) per 1-SD higher level on the natural log scale | | | | | | | |
|---|---|---|---|---|---|---|---|---|---|
| | | Standard model* | | Standard model + HbA1c | | Full model † | | Standard model excluding first 3 years of follow-up | |
| | | HR (95% CI) § | p-value ‖ | HR (95% CI) § | p-value ‖ | HR (95% CI) § | p-value ‖ | HR (95% CI) § | p-value ‖ |
| Medium LDL particles | 0.30 (0.09) mmol/L | 1.07 (1.02, 1.12) | $2.0^{-02}$ | 0.99 (0.94, 1.04) | $6.8^{-01}$ | 1.00 (0.93, 1.06) | $9.2^{-01}$ | 1.07 (1.01, 1.13) | $2.8^{-02}$ |
| Small LDL particles | 0.13 (0.03) mmol/L | 1.07 (1.02, 1.12) | $1.5^{-02}$ | 0.99 (0.94, 1.04) | $7.2^{-01}$ | 1.00 (0.93, 1.06) | $9.2^{-01}$ | 1.07 (1.02, 1.13) | $2.0^{-02}$ |
| Very large HDL particles | 0.06 (0.03) mmol/L | 0.86 (0.83, 0.89) | $4.8^{-23}$ | 0.87 (0.83, 0.90) | $7.4^{-18}$ | 0.86 (0.81, 0.91) | $7.6^{-08}$ | 0.86 (0.83, 0.89) | $1.7^{-18}$ |
| Large HDL particles | 0.23 (0.13) mmol/L | 0.77 (0.73, 0.81) | $3.8^{-35}$ | 0.81 (0.76, 0.85) | $7.1^{-20}$ | 0.83 (0.77, 0.89) | $6.1^{-09}$ | 0.77 (0.73, 0.82) | $8.2^{-29}$ |
| Medium HDL particles | 0.40 (0.10) mmol/L | 0.83 (0.77, 0.88) | $2.3^{-11}$ | 0.87 (0.82, 0.93) | $4.5^{-06}$ | 0.90 (0.84, 0.97) | $6.2^{-03}$ | 0.82 (0.76, 0.88) | $1.7^{-10}$ |
| Small HDL particles | 0.33 (0.05) mmol/L | 0.91 (0.86, 0.96) | $3.0^{-04}$ | 0.91 (0.86, 0.96) | $5.5^{-04}$ | 0.91 (0.84, 0.97) | $4.6^{-03}$ | 0.91 (0.85, 0.96) | $6.2^{-04}$ |
| **Triglyceride concentrations** | | | | | | | | | |
| Chylomicrons and extremely large VLDL particles | 0.14 (0.13) mmol/L | 1.46 (1.37, 1.55) | $2.1^{-16}$ | 1.30 (1.22, 1.38) | $1.4^{-09}$ | 1.24 (1.15, 1.34) | $3.1^{-05}$ | 1.44 (1.34, 1.53) | $3.7^{-13}$ |
| Very large VLDL particles | 0.11 (0.08) mmol/L | 1.54 (1.46, 1.61) | $2.3^{-28}$ | 1.35 (1.28, 1.43) | $4.9^{-15}$ | 1.30 (1.22, 1.39) | $2.9^{-08}$ | 1.48 (1.40, 1.56) | $5.8^{-21}$ |
| Large VLDL particles | 0.16 (0.09) mmol/L | 1.37 (1.31, 1.43) | $3.8^{-24}$ | 1.24 (1.18, 1.30) | $6.9^{-12}$ | 1.21 (1.14, 1.28) | $8.3^{-07}$ | 1.34 (1.27, 1.40) | $5.1^{-18}$ |
| Medium VLDL particles | 0.27 (0.11) mmol/L | 1.32 (1.26, 1.38) | $3.9^{-21}$ | 1.20 (1.14, 1.26) | $7.1^{-10}$ | 1.18 (1.11, 1.25) | $5.4^{-06}$ | 1.30 (1.23, 1.36) | $5.1^{-16}$ |
| Small VLDL particles | 0.16 (0.06) mmol/L | 1.40 (1.34, 1.45) | $2.3^{-31}$ | 1.27 (1.21, 1.32) | $2.3^{-16}$ | 1.26 (1.19, 1.32) | $2.3^{-10}$ | 1.37 (1.31, 1.43) | $7.7^{-24}$ |
| Very small VLDL particles | 0.07 (0.02) mmol/L | 1.40 (1.35, 1.45) | $8.9^{-38}$ | 1.27 (1.22, 1.32) | $1.4^{-19}$ | 1.28 (1.22, 1.34) | $2.4^{-14}$ | 1.38 (1.32, 1.43) | $2.1^{-29}$ |
| IDL particles | 0.10 (0.03) mmol/L | 1.37 (1.32, 1.42) | $3.8^{-37}$ | 1.25 (1.20, 1.30) | $2.9^{-18}$ | 1.27 (1.21, 1.32) | $2.7^{-14}$ | 1.36 (1.30, 1.41) | $2.1^{-29}$ |
| Large LDL particles | 0.10 (0.03) mmol/L | 1.39 (1.34, 1.43) | $3.2^{-40}$ | 1.25 (1.20, 1.30) | $5.4^{-19}$ | 1.27 (1.21, 1.32) | $2.4^{-14}$ | 1.37 (1.32, 1.42) | $7.9^{-32}$ |
| Medium LDL particles | 0.03 (0.01) mmol/L | 1.40 (1.35, 1.45) | $2.0^{-42}$ | 1.26 (1.22, 1.31) | $1.6^{-20}$ | 1.27 (1.21, 1.32) | $2.4^{-14}$ | 1.38 (1.33, 1.43) | $2.5^{-33}$ |
| Small LDL particles | 0.02 (0.01) mmol/L | 1.40 (1.35, 1.45) | $1.3^{-40}$ | 1.27 (1.22, 1.32) | $1.6^{-20}$ | 1.26 (1.20, 1.32) | $2.1^{-13}$ | 1.38 (1.32, 1.43) | $3.0^{-31}$ |
| Very large HDL particles | $7.1^{-03}$ ($2.7^{-03}$) mmol/L | 1.29 (1.24, 1.34) | $2.7^{-24}$ | 1.20 (1.15, 1.25) | $3.2^{-13}$ | 1.22 (1.16, 1.27) | $1.5^{-10}$ | 1.27 (1.21, 1.32) | $2.2^{-18}$ |
| Large HDL particles | 0.03 (0.01) mmol/L | 1.20 (1.15, 1.25) | $1.7^{-12}$ | 1.17 (1.12, 1.22) | $2.6^{-09}$ | 1.20 (1.14, 1.26) | $1.3^{-08}$ | 1.18 (1.13, 1.24) | $3.5^{-09}$ |
| Medium HDL particles | 0.05 (0.02) mmol/L | 1.34 (1.29, 1.39) | $6.2^{-27}$ | 1.27 (1.22, 1.32) | $4.8^{-18}$ | 1.27 (1.21, 1.34) | $9.1^{-13}$ | 1.31 (1.25, 1.37) | $8.6^{-20}$ |
| Small HDL particles | 0.05 (0.02) mmol/L | 1.51 (1.46, 1.57) | $4.3^{-42}$ | 1.37 (1.31, 1.43) | $1.6^{-24}$ | 1.35 (1.28, 1.42) | $3.1^{-15}$ | 1.48 (1.41, 1.54) | $8.6^{-32}$ |
| **Phospholipid concentrations** | | | | | | | | | |
| Chylomicrons and extremely large VLDL particles | 0.04 (0.03) mmol/L | 1.54 (1.45, 1.64) | $1.9^{-18}$ | 1.37 (1.28, 1.46) | $2.1^{-11}$ | 1.33 (1.22, 1.44) | $1.0^{-06}$ | 1.48 (1.38, 1.58) | $9.3^{-14}$ |
| Very large VLDL particles | 0.04 (0.03) mmol/L | 1.58 (1.49, 1.68) | $1.6^{-20}$ | 1.36 (1.27, 1.45) | $1.5^{-10}$ | 1.30 (1.19, 1.41) | $1.4^{-05}$ | 1.52 (1.41, 1.62) | $5.1^{-15}$ |
| Large VLDL particles | 0.07 (0.04) mmol/L | 1.60 (1.50, 1.70) | $8.1^{-20}$ | 1.36 (1.27, 1.46) | $7.6^{-10}$ | 1.30 (1.18, 1.41) | $4.2^{-05}$ | 1.53 (1.42, 1.64) | $2.1^{-14}$ |
| Medium VLDL particles | 0.13 (0.05) mmol/L | 1.12 (1.06, 1.17) | $1.1^{-04}$ | 1.03 (0.98, 1.09) | $3.1^{-01}$ | 1.03 (0.97, 1.10) | $4.0^{-01}$ | 1.11 (1.05, 1.17) | $8.6^{-04}$ |
| Small VLDL particles | 0.10 (0.03) mmol/L | 1.15 (1.09, 1.20) | $1.6^{-06}$ | 1.05 (0.99, 1.10) | $1.1^{-01}$ | 1.06 (0.99, 1.12) | $1.6^{-01}$ | 1.14 (1.08, 1.20) | $2.1^{-05}$ |

#### Webtable 1. Distribution of metabolic biomarkers and their associations with incident type 2 diabetes

| Metabolic biomarker | Mean (SD) | Hazard ratio (95% CI) per 1-SD higher level on the natural log scale | | | | | | | |
|---|---|---|---|---|---|---|---|---|---|
| | | Standard model* | | Standard model + HbA1c | | Full model † | | Standard model excluding first 3 years of follow-up | |
| | | HR (95% CI) § | p-value ‖ | HR (95% CI) § | p-value ‖ | HR (95% CI) § | p-value ‖ | HR (95% CI) § | p-value ‖ |
| Very small VLDL particles | 0.10 (0.03) mmol/L | 1.19 (1.14, 1.25) | $4.3^{-11}$ | 1.09 (1.04, 1.14) | $1.3^{-03}$ | 1.12 (1.06, 1.18) | $7.7^{-04}$ | 1.19 (1.13, 1.24) | $3.2^{-09}$ |
| IDL particles | 0.29 (0.07) mmol/L | 0.96 (0.91, 1.01) | $1.0^{-01}$ | 0.91 (0.86, 0.96) | $2.8^{-04}$ | 0.94 (0.88, 1.00) | $7.8^{-02}$ | 0.96 (0.91, 1.02) | $2.1^{-01}$ |
| Large LDL particles | 0.35 (0.08) mmol/L | 0.95 (0.90, 1.00) | $5.5^{-02}$ | 0.90 (0.85, 0.95) | $6.7^{-05}$ | 0.92 (0.86, 0.98) | $1.3^{-02}$ | 0.96 (0.90, 1.01) | $1.3^{-01}$ |
| Medium LDL particles | 0.16 (0.04) mmol/L | 1.04 (0.99, 1.10) | $1.1^{-01}$ | 0.97 (0.92, 1.02) | $2.5^{-01}$ | 0.98 (0.91, 1.04) | $5.4^{-01}$ | 1.05 (0.99, 1.10) | $1.2^{-01}$ |
| Small LDL particles | 0.09 (0.02) mmol/L | 1.05 (1.01, 1.10) | $4.2^{-02}$ | 0.98 (0.93, 1.02) | $3.6^{-01}$ | 0.99 (0.93, 1.05) | $8.4^{-01}$ | 1.06 (1.01, 1.11) | $4.2^{-02}$ |
| Very large HDL particles | 0.08 (0.04) mmol/L | 0.91 (0.87, 0.95) | $8.8^{-06}$ | 0.92 (0.88, 0.96) | $2.6^{-04}$ | 0.94 (0.88, 1.00) | $7.0^{-02}$ | 0.91 (0.87, 0.96) | $6.5^{-05}$ |
| Large HDL particles | 0.32 (0.15) mmol/L | 0.89 (0.85, 0.93) | $7.5^{-09}$ | 0.90 (0.85, 0.95) | $4.4^{-05}$ | 0.94 (0.87, 1.01) | $1.0^{-01}$ | 0.89 (0.84, 0.93) | $6.1^{-08}$ |
| Medium HDL particles | 0.48 (0.10) mmol/L | 0.99 (0.94, 1.05) | $8.1^{-01}$ | 1.01 (0.96, 1.06) | $7.0^{-01}$ | 1.04 (0.98, 1.11) | $2.4^{-01}$ | 0.98 (0.92, 1.03) | $4.4^{-01}$ |
| Small HDL particles | 0.65 (0.09) mmol/L | 1.11 (1.06, 1.16) | $1.6^{-04}$ | 1.09 (1.03, 1.14) | $2.9^{-03}$ | 1.09 (1.03, 1.16) | $9.4^{-03}$ | 1.08 (1.03, 1.14) | $5.5^{-03}$ |
| **Total lipid concentrations** | | | | | | | | | |
| Chylomicrons and extremely large VLDL particles | 0.23 (0.21) mmol/L | 1.46 (1.39, 1.53) | $1.3^{-25}$ | 1.32 (1.25, 1.39) | $9.3^{-15}$ | 1.28 (1.20, 1.36) | $1.7^{-08}$ | 1.42 (1.35, 1.50) | $9.7^{-20}$ |
| Very large VLDL particles | 0.20 (0.13) mmol/L | 1.49 (1.42, 1.56) | $2.1^{-27}$ | 1.32 (1.25, 1.39) | $3.0^{-14}$ | 1.28 (1.20, 1.37) | $5.1^{-08}$ | 1.45 (1.37, 1.52) | $1.3^{-20}$ |
| Large VLDL particles | 0.33 (0.17) mmol/L | 1.39 (1.32, 1.45) | $5.6^{-23}$ | 1.24 (1.18, 1.31) | $3.6^{-11}$ | 1.22 (1.14, 1.29) | $2.1^{-06}$ | 1.35 (1.29, 1.42) | $2.6^{-17}$ |
| Medium VLDL particles | 0.57 (0.20) mmol/L | 1.18 (1.13, 1.24) | $7.7^{-09}$ | 1.08 (1.03, 1.14) | $7.8^{-03}$ | 1.08 (1.01, 1.15) | $4.5^{-02}$ | 1.17 (1.11, 1.23) | $6.5^{-07}$ |
| Small VLDL particles | 0.41 (0.13) mmol/L | 1.27 (1.21, 1.33) | $2.9^{-16}$ | 1.15 (1.09, 1.20) | $3.6^{-06}$ | 1.15 (1.08, 1.22) | $2.1^{-04}$ | 1.26 (1.19, 1.32) | $8.1^{-13}$ |
| Very small VLDL particles | 0.36 (0.09) mmol/L | 1.17 (1.11, 1.22) | $1.2^{-08}$ | 1.07 (1.02, 1.12) | $1.4^{-02}$ | 1.10 (1.04, 1.17) | $4.5^{-03}$ | 1.16 (1.11, 1.22) | $3.4^{-07}$ |
| IDL particles | 1.22 (0.29) mmol/L | 0.95 (0.90, 1.00) | $4.6^{-02}$ | 0.90 (0.85, 0.95) | $7.2^{-05}$ | 0.93 (0.87, 0.99) | $4.4^{-02}$ | 0.95 (0.90, 1.01) | $9.8^{-02}$ |
| Large LDL particles | 1.56 (0.37) mmol/L | 0.98 (0.93, 1.02) | $3.4^{-01}$ | 0.92 (0.87, 0.97) | $1.8^{-03}$ | 0.94 (0.88, 1.00) | $8.1^{-02}$ | 0.98 (0.93, 1.03) | $4.9^{-01}$ |
| Medium LDL particles | 0.61 (0.16) mmol/L | 1.06 (1.01, 1.11) | $4.1^{-02}$ | 0.98 (0.93, 1.03) | $4.6^{-01}$ | 0.99 (0.93, 1.05) | $7.9^{-01}$ | 1.06 (1.00, 1.12) | $4.9^{-02}$ |
| Small LDL particles | 0.28 (0.06) mmol/L | 1.07 (1.02, 1.12) | $1.3^{-02}$ | 0.99 (0.94, 1.04) | $6.1^{-01}$ | 1.00 (0.94, 1.06) | $9.2^{-01}$ | 1.07 (1.02, 1.13) | $1.8^{-02}$ |
| Very large HDL particles | 0.17 (0.08) mmol/L | 0.84 (0.78, 0.89) | $3.1^{-09}$ | 0.85 (0.79, 0.91) | $2.3^{-07}$ | 0.89 (0.82, 0.97) | $7.6^{-03}$ | 0.84 (0.78, 0.90) | $2.1^{-07}$ |
| Large HDL particles | 0.64 (0.32) mmol/L | 0.80 (0.74, 0.85) | $1.2^{-13}$ | 0.84 (0.78, 0.90) | $3.2^{-08}$ | 0.89 (0.81, 0.96) | $4.3^{-03}$ | 0.79 (0.73, 0.86) | $6.8^{-12}$ |
| Medium HDL particles | 1.02 (0.22) mmol/L | 0.94 (0.89, 1.00) | $4.6^{-02}$ | 0.97 (0.92, 1.03) | $3.3^{-01}$ | 1.01 (0.94, 1.07) | $9.0^{-01}$ | 0.93 (0.87, 0.99) | $2.0^{-02}$ |
| Small HDL particles | 1.15 (0.16) mmol/L | 1.09 (1.04, 1.14) | $1.5^{-03}$ | 1.06 (1.01, 1.12) | $3.1^{-02}$ | 1.07 (1.01, 1.13) | $6.5^{-02}$ | 1.07 (1.02, 1.13) | $2.0^{-02}$ |
| **Lipoprotein particle sizes** | | | | | | | | | |
| VLDL particles | 38.6 (1.3) nm | 1.36 (1.30, 1.41) | $3.1^{-24}$ | 1.27 (1.21, 1.33) | $3.0^{-15}$ | 1.21 (1.14, 1.28) | $5.4^{-07}$ | 1.32 (1.26, 1.39) | $5.4^{-18}$ |
| LDL particles | 23.9 (0.1) nm | 0.82 (0.78, 0.87) | $3.0^{-17}$ | 0.86 (0.81, 0.90) | $3.9^{-11}$ | 0.87 (0.82, 0.93) | $1.8^{-06}$ | 0.83 (0.78, 0.88) | $7.2^{-14}$ |
| HDL particles | 9.64 (0.21) nm | 0.78 (0.71, 0.85) | $2.7^{-11}$ | 0.84 (0.77, 0.92) | $6.7^{-06}$ | 0.91 (0.82, 1.00) | $4.8^{-02}$ | 0.78 (0.70, 0.86) | $8.5^{-10}$ |

#### Webtable 1. Distribution of metabolic biomarkers and their associations with incident type 2 diabetes

| Metabolic biomarker | Mean (SD) | Hazard ratio (95% CI) per 1-SD higher level on the natural log scale | | | | | | | |
|---|---|---|---|---|---|---|---|---|---|
| | | Standard model* | | Standard model + HbA1c | | Full model † | | Standard model excluding first 3 years of follow-up | |
| | | HR (95% CI) § | p-value \|\| | HR (95% CI) § | p-value \|\| | HR (95% CI) § | p-value \|\| | HR (95% CI) § | p-value \|\| |
| **Apolipoproteins** | | | | | | | | | |
| Apolipoprotein A1 | 1.44 (0.24) g/L | 0.91 (0.85, 0.97) | $1.4^{-03}$ | 0.93 (0.88, 0.99) | $3.4^{-02}$ | 0.98 (0.91, 1.05) | $5.9^{-01}$ | 0.90 (0.84, 0.96) | $8.7^{-04}$ |
| Apolipoprotein B | 0.84 (0.20) g/L | 1.04 (0.99, 1.09) | $1.5^{-01}$ | 0.97 (0.92, 1.02) | $2.1^{-01}$ | 0.98 (0.92, 1.04) | $5.9^{-01}$ | 1.04 (0.99, 1.10) | $1.5^{-01}$ |
| Apolipoprotein B to Apolipoprotein A1 ratio | 0.60 (0.17) | 1.09 (1.04, 1.15) | $2.3^{-03}$ | 1.00 (0.95, 1.06) | $9.4^{-01}$ | 0.99 (0.93, 1.06) | $8.7^{-01}$ | 1.10 (1.04, 1.16) | $1.9^{-03}$ |
| **Fatty acids** | | | | | | | | | |
| Polyunsaturated | 4.98 (0.80) mmol/L | 1.04 (0.99, 1.09) | $1.6^{-01}$ | 0.98 (0.93, 1.03) | $5.0^{-01}$ | 1.00 (0.94, 1.06) | $9.2^{-01}$ | 1.04 (0.98, 1.09) | $2.3^{-01}$ |
| Monounsaturated | 2.82 (0.82) mmol/L | 1.33 (1.28, 1.38) | $1.2^{-28}$ | 1.23 (1.18, 1.28) | $1.2^{-14}$ | 1.21 (1.15, 1.27) | $2.6^{-09}$ | 1.31 (1.26, 1.37) | $4.7^{-22}$ |
| Saturated | 4.04 (0.95) mmol/L | 1.31 (1.27, 1.36) | $1.2^{-28}$ | 1.20 (1.15, 1.25) | $1.8^{-13}$ | 1.21 (1.15, 1.26) | $7.6^{-10}$ | 1.29 (1.24, 1.34) | $9.6^{-22}$ |
| Docosahexaenoic acid | 0.23 (0.08) mmol/L | 0.90 (0.85, 0.94) | $8.0^{-06}$ | 0.91 (0.86, 0.96) | $1.3^{-04}$ | 0.93 (0.87, 0.99) | $4.2^{-02}$ | 0.90 (0.85, 0.95) | $4.8^{-05}$ |
| Linoleic acid | 3.41 (0.68) mmol/L | 1.02 (0.97, 1.07) | $3.9^{-01}$ | 0.96 (0.91, 1.01) | $1.3^{-01}$ | 0.97 (0.91, 1.03) | $3.8^{-01}$ | 1.02 (0.97, 1.08) | $4.4^{-01}$ |
| Omega-3 | 0.53 (0.22) mmol/L | 1.03 (0.98, 1.08) | $2.8^{-01}$ | 1.01 (0.96, 1.06) | $7.0^{-01}$ | 1.04 (0.98, 1.10) | $3.1^{-01}$ | 1.02 (0.97, 1.08) | $4.4^{-01}$ |
| Omega-6 | 4.45 (0.68) mmol/L | 1.04 (0.99, 1.09) | $1.7^{-01}$ | 0.98 (0.93, 1.03) | $3.7^{-01}$ | 0.99 (0.92, 1.05) | $6.9^{-01}$ | 1.04 (0.98, 1.09) | $2.1^{-01}$ |
| Total | 11.8 (2.4) mmol/L | 1.25 (1.20, 1.30) | $7.9^{-19}$ | 1.15 (1.10, 1.20) | $4.3^{-08}$ | 1.15 (1.10, 1.21) | $5.4^{-06}$ | 1.23 (1.18, 1.29) | $2.1^{-14}$ |
| Polyunsaturated to total fatty acids ratio | 42.5 (3.8) % | 0.70 (0.66, 0.74) | $7.4^{-60}$ | 0.76 (0.71, 0.80) | $9.3^{-36}$ | 0.76 (0.71, 0.81) | $1.0^{-22}$ | 0.72 (0.67, 0.76) | $3.3^{-45}$ |
| Monounsaturated to total fatty acids ratio | 23.5 (2.7) % | 1.42 (1.37, 1.48) | $7.3^{-36}$ | 1.33 (1.28, 1.39) | $2.0^{-23}$ | 1.30 (1.23, 1.37) | $2.1^{-13}$ | 1.40 (1.34, 1.46) | $3.4^{-28}$ |
| Saturated to total fatty acids ratio | 34.0 (2.0) % | 1.36 (1.31, 1.40) | $7.1^{-39}$ | 1.26 (1.22, 1.31) | $3.0^{-23}$ | 1.28 (1.23, 1.34) | $5.8^{-18}$ | 1.33 (1.28, 1.38) | $2.1^{-29}$ |
| Docosahexaenoic to total fatty acids ratio | 2.01 (0.68) % | 0.81 (0.76, 0.85) | $3.0^{-21}$ | 0.85 (0.81, 0.90) | $7.0^{-12}$ | 0.86 (0.80, 0.92) | $1.6^{-06}$ | 0.81 (0.76, 0.86) | $4.2^{-17}$ |
| Linoleic to total fatty acids ratio | 29.0 (3.4) % | 0.74 (0.69, 0.78) | $3.8^{-41}$ | 0.76 (0.72, 0.81) | $1.1^{-31}$ | 0.77 (0.72, 0.82) | $2.2^{-20}$ | 0.75 (0.70, 0.80) | $6.6^{-31}$ |
| Omega-3 to total fatty acids ratio | 4.42 (1.56) % | 0.92 (0.87, 0.96) | $6.8^{-04}$ | 0.94 (0.89, 0.99) | $1.4^{-02}$ | 0.95 (0.88, 1.02) | $2.0^{-01}$ | 0.92 (0.86, 0.97) | $1.8^{-03}$ |
| Omega-6 to total fatty acids ratio | 38.1 (3.6) % | 0.72 (0.68, 0.77) | $1.8^{-51}$ | 0.77 (0.73, 0.81) | $6.9^{-32}$ | 0.78 (0.73, 0.83) | $1.3^{-20}$ | 0.74 (0.69, 0.78) | $3.6^{-38}$ |
| **Other lipids** | | | | | | | | | |
| Total cholines | 2.53 (0.41) mmol/L | 1.04 (0.99, 1.10) | $1.3^{-01}$ | 1.01 (0.95, 1.06) | $8.5^{-01}$ | 1.04 (0.98, 1.11) | $2.4^{-01}$ | 1.03 (0.98, 1.09) | $2.7^{-01}$ |
| Phosphatidylcholines | 2.08 (0.37) mmol/L | 1.07 (1.02, 1.12) | $1.7^{-02}$ | 1.04 (0.99, 1.09) | $2.1^{-01}$ | 1.08 (1.02, 1.15) | $2.9^{-02}$ | 1.05 (1.00, 1.11) | $8.6^{-02}$ |
| Sphingomyelins | 0.44 (0.07) mmol/L | 0.92 (0.87, 0.97) | $1.8^{-03}$ | 0.88 (0.83, 0.93) | $3.6^{-06}$ | 0.91 (0.85, 0.98) | $8.6^{-03}$ | 0.93 (0.87, 0.99) | $1.5^{-02}$ |
| Phosphoglycerides | 2.25 (0.40) mmol/L | 1.09 (1.04, 1.15) | $9.9^{-04}$ | 1.05 (1.00, 1.10) | $9.4^{-02}$ | 1.09 (1.02, 1.15) | $1.9^{-02}$ | 1.08 (1.02, 1.14) | $9.8^{-03}$ |
| **Glycolysis-related metabolites** | | | | | | | | | |
| Lactate | 3.81 (1.09) mmol/L | 1.14 (1.08, 1.19) | $3.9^{-06}$ | 1.11 (1.06, 1.16) | $1.2^{-04}$ | 1.10 (1.04, 1.17) | $4.1^{-03}$ | 1.12 (1.07, 1.18) | $9.8^{-05}$ |
| Citrate | 0.06 (0.01) mmol/L | 1.07 (1.01, 1.12) | $2.0^{-02}$ | 1.04 (0.98, 1.09) | $2.3^{-01}$ | 1.05 (0.99, 1.11) | $1.7^{-01}$ | 1.08 (1.02, 1.13) | $1.3^{-02}$ |
| Glucose | 3.57 (1.14) mmol/L | 1.68 (1.63, 1.73) | $1.2^{-88}$ | 1.30 (1.25, 1.34) | $2.9^{-24}$ | 1.28 (1.23, 1.34) | $1.0^{-15}$ | 1.60 (1.54, 1.65) | $1.2^{-60}$ |

#### Webtable 1. Distribution of metabolic biomarkers and their associations with incident type 2 diabetes

| Metabolic biomarker | Mean (SD) | Hazard ratio (95% CI) per 1-SD higher level on the natural log scale | | | | | | | |
|---|---|---|---|---|---|---|---|---|---|
| | | Standard model* | | Standard model + HbA1c | | Full model † | | Standard model excluding first 3 years of follow-up | |
| | | HR (95% CI) § | p-value \|\| | HR (95% CI) § | p-value \|\| | HR (95% CI) § | p-value \|\| | HR (95% CI) § | p-value \|\| |
| **Amino acids** | | | | | | | | | |
| Alanine | 0.30 (0.08) mmol/L | 1.29 (1.23, 1.35) | $4.4^{-16}$ | 1.17 (1.11, 1.23) | $6.0^{-07}$ | 1.15 (1.07, 1.22) | $5.7^{-04}$ | 1.25 (1.19, 1.32) | $3.4^{-11}$ |
| Glutamine | 0.53 (0.08) mmol/L | 0.89 (0.84, 0.94) | $6.3^{-06}$ | 0.93 (0.88, 0.98) | $3.7^{-03}$ | 0.92 (0.86, 0.98) | $7.7^{-03}$ | 0.91 (0.86, 0.96) | $4.8^{-04}$ |
| Histidine | 0.06 (0.01) mmol/L | 0.95 (0.90, 1.00) | $5.0^{-02}$ | 0.96 (0.91, 1.01) | $1.5^{-01}$ | 0.98 (0.92, 1.04) | $6.1^{-01}$ | 0.94 (0.89, 1.00) | $3.6^{-02}$ |
| Glycine | 0.16 (0.06) mmol/L | 0.82 (0.78, 0.86) | $7.3^{-22}$ | 0.86 (0.81, 0.90) | $4.8^{-11}$ | 0.85 (0.79, 0.90) | $8.1^{-09}$ | 0.82 (0.78, 0.87) | $3.5^{-16}$ |
| Isoleucine | 0.05 (0.02) mmol/L | 1.19 (1.14, 1.24) | $5.6^{-11}$ | 1.13 (1.07, 1.18) | $1.3^{-05}$ | 1.11 (1.04, 1.17) | $2.9^{-03}$ | 1.19 (1.14, 1.25) | $7.7^{-10}$ |
| Leucine | 0.10 (0.03) mmol/L | 1.19 (1.14, 1.24) | $1.5^{-10}$ | 1.14 (1.09, 1.19) | $1.4^{-06}$ | 1.12 (1.06, 1.18) | $1.1^{-03}$ | 1.19 (1.13, 1.24) | $4.5^{-09}$ |
| Valine | 0.20 (0.04) mmol/L | 1.31 (1.26, 1.37) | $4.9^{-24}$ | 1.21 (1.16, 1.26) | $8.2^{-12}$ | 1.19 (1.13, 1.26) | $3.0^{-07}$ | 1.31 (1.26, 1.37) | $1.7^{-20}$ |
| Phenylalanine | 0.05 (0.01) mmol/L | 1.11 (1.06, 1.16) | $4.7^{-05}$ | 1.05 (1.00, 1.10) | $8.6^{-02}$ | 1.06 (1.00, 1.12) | $6.9^{-02}$ | 1.12 (1.06, 1.17) | $7.9^{-05}$ |
| Tyrosine | 0.06 (0.01) mmol/L | 1.33 (1.27, 1.38) | $1.0^{-26}$ | 1.19 (1.14, 1.24) | $2.8^{-10}$ | 1.21 (1.15, 1.28) | $7.0^{-09}$ | 1.32 (1.26, 1.37) | $1.1^{-21}$ |
| **Ketone bodies** | | | | | | | | | |
| Acetate mmol/l | 0.02 (0.03) mmol/L | 0.95 (0.91, 1.00) | $3.8^{-02}$ | 1.00 (0.96, 1.05) | $8.7^{-01}$ | 1.02 (0.96, 1.07) | $6.3^{-01}$ | 0.95 (0.90, 1.00) | $7.4^{-02}$ |
| Acetoacetate mmol/l | 0.01 (0.01) mmol/L | 1.04 (0.98, 1.09) | $2.1^{-01}$ | 1.03 (0.98, 1.09) | $3.0^{-01}$ | 1.03 (0.97, 1.10) | $4.4^{-01}$ | 1.05 (1.00, 1.11) | $8.2^{-02}$ |
| 3-Hydroxybutyrate mmol/l | 0.01 (0.01) mmol/L | 0.88 (0.83, 0.94) | $1.3^{-05}$ | 0.95 (0.90, 1.01) | $1.1^{-01}$ | 0.98 (0.92, 1.05) | $6.6^{-01}$ | 0.90 (0.84, 0.96) | $6.2^{-04}$ |
| Acetone | 0.08 (0.03) mmol/L | 1.06 (1.00, 1.11) | $5.7^{-02}$ | 1.03 (0.97, 1.08) | $4.1^{-01}$ | 1.01 (0.94, 1.07) | $8.5^{-01}$ | 1.06 (1.00, 1.12) | $5.2^{-02}$ |
| Pyruvate | 0.06 (0.06) mmol/L | 0.98 (0.92, 1.03) | $4.0^{-01}$ | 0.98 (0.92, 1.04) | $5.8^{-01}$ | 0.99 (0.92, 1.06) | $7.6^{-01}$ | 0.99 (0.93, 1.05) | $6.7^{-01}$ |
| **Fluid balance** | | | | | | | | | |
| Albumin | 38.9 (3.3) g/L | 0.99 (0.94, 1.04) | $6.5^{-01}$ | 1.08 (1.03, 1.13) | $6.8^{-03}$ | 1.11 (1.05, 1.17) | $1.4^{-03}$ | 0.98 (0.93, 1.04) | $5.9^{-01}$ |
| Creatinine | 0.07 (0.02) mmol/L | 1.09 (1.04, 1.15) | $2.9^{-03}$ | 1.06 (1.01, 1.12) | $4.9^{-02}$ | 1.04 (0.97, 1.11) | $3.6^{-01}$ | 1.09 (1.03, 1.15) | $9.3^{-03}$ |
| **Inflammation** | | | | | | | | | |
| Glycoprotein acetyls | 0.79 (0.12) mmol/L | 1.32 (1.27, 1.38) | $3.8^{-23}$ | 1.09 (1.04, 1.15) | $2.9^{-03}$ | 1.06 (1.00, 1.13) | $1.1^{-01}$ | 1.33 (1.27, 1.39) | $8.4^{-21}$ |

* Standard model: Stratified by age-at-risk and sex and adjusted for assessment centre, Townsend deprivation index, smoking, alcohol drinking, body mass index, waist-to-hip ratio, fasting duration and spectrometer
† Full model: Standard model additionally adjusted for HbA1c, ethnicity, parental history of diabetes, physical activity, and intakes of whole grains, refined grains, fruit, vegetables, cheese, unprocessed red meat, processed meat, non-oily fish, oily fish, type of spread, coffee (regular and decaffeinated), tea, and dietary supplements
§ HR per 1-SD higher metabolic biomarker on the natural log scale
|| Controlled for false discovery rate

**Webtable 2. Diagnosis and medication codes for assessment of type 2 diabetes status in primary and secondary healthcare and death registry records and UK Biobank verbal interview**

| Code format | Codes |
|---|---|
| ***Diagnosis codes*** | |
| **ICD10** | E11 |
| **Read v2** | C1001, C1011, C1021, C1031, C1041, C1051, C1061, C1071, C1074, C109., C1090, C1091, C1092, C1093, C1094, C1095, C1096, C1097, C1099, C109A, C109B, C109C, C109D, C109E, C109F, C109G, C109H, C109J, C109K, C10F., C10F0, C10F1, C10F2, C10F3, C10F4, C10F5, C10F6, C10F7, C10F9, C10FA, C10FB, C10FC, C10FD, C10FE, C10FF, C10FG, C10FH, C10FJ, C10FK, C10FL, C10FM, C10FN, C10FP, C10FQ, C10FR, C10P1, C10y1, C10z1, L1806, L180B, |
| **Read CTv3** | 66A3., C1001, C1011, C1021, C1031, C1041, C1051, C1061, C1071, C1074, C109., C1090, C1091, C1092, C1093, C1094, C1095, C1096, C1097, C10y1, C10z1, L1806, X40J5, X40J6, Xaagf, XacsX, XacsY, XaELQ, XaEnp, XaEnq, XaF05, XaFmA, XaFn7, XaFn8, aFn8, XaFn9, XaFWI, XaIfG, XaIfI, XaIzQ, XaIzR, XaJQp, XaKyX, XaXZR, XE10F, XE12A, XE12A, XE12A, XE12A, XM19j |
| ***Medication codes**** | |
| **Read v2** | f1..., , f11.., f111., f112., f12.., f121., f122., f123., f124., f125., f126., f127., f128., f129., f12A., f12a., f12B., f12C., f12D., f12d., f12E., f12e., f12F., f12g., f12G., f12H., f12I., f12J., f12K., f12L., f12M., f12Q., f12R., f12s., f12S., f12T., f12U., f12V., f12W., f12X., f12y., f12Y., f12z., f12Z., f13.., f131., f132., f133., f134., f135., f136., f137., f138., f139., f13A., f14.., f141., f142., f143., f144., f145., f146., f14w., f14x., f14y., f14z., f15.., f151., f152., f153., f154., f155., f15x., f15y., f15z., f2..., f21.., f211., f212., f22.., f221., f222., f223., f224., f225., f226., f227., f228., f23.., f231., f24.., f241., f242., f25.., f251., f252., f253., f254., f255., f256., f257., f258., f259., f25a., f25A., f25b., f25B., f25c., f25C., f25d., f25D., f25e., f25E., f25f., f25F., f25g., f25G., f25h., f25H., f25i., f25j., f25k., f25l., f25m., f25n., f25o., f25p., f25q., f25r., f25s., f25t., f25u., f25v., f25W., f25w., f25X., f25x., f25Y., f25y., f25z., f26.., f261., f262., f27.., f271., f272., f273., f274., f275., f276., f277., f278., f279., f27A., f27a., f27b., f27B., f27C., f27c., f27D., f27d., f27E., f27e., f27f., f27F., f27g., f27G., f27h., f27H., f27i., f27I., f27J., f27j., f27k., f27K., f27l., f27L., f27m., f27M., f27n., f27N., f27o., f27O., f27P., f27p., f27Q., f27q., f27R., f27r., f27S., f27s., f27T., f27t., f27u., f27v., f27V., f27W., f27w., f27x., f27X., f27Y., f27y., f27z., f27Z., f28.., f281., f282., f283., f284., f285., f286., f287., f288., f289., f28A., f28B., f28C., f28D., f28E., f28F., f29.., f291., f292., f293., f294., f295., f296., f297., f298., f299., f29A., f29B., f29C., f2A.., f2A1., f2A2., f2A3., f2Ax., f2Ay., f2Az., f2B.., f2B1., f2B2., f2B3., f2B4., f2B5., f2B6., f2C.., f2C1., f2C2., f3..., f31.., f311., f31z., f32.., f321., f322., f323., f324., f325., f33.., f331., f332., f333., f334., f335., f336., f337., f338., f339., f33a., f33b., f33c., f33d., f33e., f33f., f33g., f34.., f341., f34z., f35.., f351., f352., f353., f354., f355., f356., f357., f358., f359., f35A., f35B., 35C., f35D., f35w., f35x., f35y., f35z., f36.., f361., f362., f363., f364., f36y., f36z., f37.., f371., f37z., f38.., f381., f38z., f39.., f391., f392., f39y., f39z., f3A.., f3a.., f3A1., f3a1., f3A2., f3a2., f3a3., f3A3., f3a4., f3A4., f3A5., f3A6., f3A7., f3A8., f3A9., f3AA., f3AB., f3AC., f4..., f41.., f411., f412., f413., f414., f415., f416., f417., f418., f419., f41A., f41B., f41C., f41D., f41E., f41F., f41G., f41H., f41I., f41J., f41s., f41t., |

| | |
|---|---|
| **Read v2 continued…** | f41u., f41v., f41w., f41x., f41y., f41z., ft1.., ft11., ft12., ft13., ft14., ft2.., ft21., ft22., ft23., ft24., ft25., ft26., ft3.., ft31., ft32., ft33., ft34., ft35., ft36., ft37., ft38., ft39., ft4.., ft41., ft42., ft43., ft44., ft45., ft46., ft4u., ft4v., ft4w., ft4x., ft4y., ft4z., ft5.., ft51., ft52., ft53., ft54., ft55., ft56., ft5x., ft5y., ft5z., ft6.., ft61., ft62., ft63., ft6x., ft6y., ft6z., ft7.., ft71., ft7z., ft8.., ft81., ft82., ft83., ft8x., ft8y., ft8z., ft9.., ft91., ft92., ft93., ft94., ft95., ft9x., ft9y., ft9z., fta.., fta1., ftaZ., ftb.., ftb1., ftb2., ftby., ftbz., ftc.., ftc1., ftc2., ftd.., ftd1., ftd2., ftdy., ftdz., fte.., fte1., ftez., ftf.., ftf1., ftf2., ftg.., ftg1., ftg2., ftg3., ftg4., fth.., fth1., fth2., fth3., fth4., fti.., fti1., fti2., fti3., fti4., ftj.., ftj1., ftj2., ftj3., ftj4., ftj5., ftj6. ftk.., ftk1., ftk2., ftk3., ftk4., ftk5., ftk6., ftl.., ftl1., ftl2., ftm.., ftm1., ftm2., ftm3., ftm4., ftn.., ftn1., ftn2., ftn3., ftn4., fto.., fto1., fto2., fto3., fto4., ftp.., ftp1., ftp2., ftp3., ftp4., ftq.., ftq1., ftq2., ftq3., ftq4., ftq5., ftq6., ftq7., ftq8., ftr.., ftr1., ftr2., ftr3., ftr4., ftr5., ftr6., ftr7., ftr8., fts.., fts1., fts2., fw..., fw1.., fw11., fw12., fw13., fw14., fw15., fw16., fw2.., fw21., fw22., pm1e. |
| **British National Formulary** | 0601011A0, 0601011A0, 0601011A0, 0601011A0, 0601011P0, 0601011Q0, 0601011R0, 060101200, 0601012C0, 0601012D0, 0601012F0, 0601012G0, 0601012L0, 0601012N0, 0601012S0, 0601012U0, 0601012V0, 0601012W0, 0601012X0, 0601012Z0, 0601021A0, 0601021B0, 0601021E0, 0601021H0, 0601021K0, 0601021M0, 0601021P0, 0601021R0, 0601021T0, 0601021V0, 0601021X0, 0601022B0, 0601022P0, 0601023A0, 0601023AA, 0601023AB, 0601023AC, 0601023AD, 0601023AE, 0601023AF, 0601023AG, 0601023AH, 0601023AI, 0601023AJ, 0601023AK, 0601023AL, 0601023AM, 0601023AN, 0601023AP, 0601023AQ, 0601023AR, 0601023AS, 0601023AU, 0601023AV, 0601023AW, 0601023AX, 0601023B0, 0601023M0, 0601023R0, 0601023S0, 0601023T0, 0601023U0, 0601023V0, 0601023W0, 0601023X0, 0601023Y0,0601023Z0 |
| **UKB verbal interview** | 1140857494, 1140857496, 1140857500, 1140857502, 1140857506, 1140857584, 1140857586, 1140857590, 1140868902, 1140868908, 1140874646, 1140874650, 1140874652, 1140874658, 1140874660, 1140874664, 1140874666, 1140874674, 1140874678, 1140874680, 1140874686, 1140874690, 1140874706, 1140874712, 1140874716, 1140874718, 1140874724, 1140874726, 1140874728, 1140874732, 1140874736, 1140874740, 1140874744, 1140874746, 1140882964, 1140883066, 1140884600, 1140910564, 1140910566, 1140910818, 1140921964, 1141152590, 1141153254, 1141153262, 1141156984, 1141157284, 1141168660, 1141168668, 1141169504, 1141171508, 1141171646, 1141171652, 1141173786, 1141173882, 1141177600, 1141177606, 1141189090, 1141189094 |

*Participants for whom receiving metformin (self-reported use or recorded in primary care data) was the only evidence of a diagnosis of diabetes were not considered to have diabetes given other indications for its use. Participants for whom receiving insulin (self-reported use or a prescription recorded in primary care data) was the only evidence of a diagnosis of diabetes were not considered to have type 2 diabetes.

**Webtable 3. Baseline characteristics of UK Biobank participants with and without NMR-metabolomics profiling**

| Baseline characteristics* | Included in NMR-metabolomics profiling | |
|---|---|---|
| | Yes | No |
| **No. of participants** | 118036 | 384454 |
| **Age, sex and socioeconomic factors** | | |
| Mean age (SD), years | 56.5 (8.1) | 56.5 (8.1) |
| Women, % | 54 | 54 |
| Townsend Deprivation Index (SD)† | 0.0 (1.0) | 0.0 (1.0) |
| **Lifestyle factors** | | |
| Smoking, % | | |
| Never or occasional | 57 | 58 |
| Previous | 35 | 34 |
| Current regular | 8 | 8 |
| Alcohol drinking, % | | |
| Never or occasional | 27 | 27 |
| Previous | 4 | 4 |
| Current regular | 69 | 69 |
| **Anthropometry, mean (SD)** | | |
| BMI, kg/$m^2$ | 27.4 (4.8) | 27.4 (4.8) |
| WC, cm | 90 (13) | 90 (14) |
| HC, cm | 103 (9) | 103 (9) |
| WHR | 0.87 (0.09) | 0.87 (0.09) |
| **Parental history of diabetes, %** | 20 | 20 |
| **Mean fasting time (SD), hours** | 3.8 (2.4) | 3.8 (2.5) |

*Standardised to age and sex structure of the study population

†Standardised Townsend Deprivation Index, higher scores represent higher levels of deprivation

BMI=body mass index; HC=hip circumference; WC=waist circumference; WHR=waist-to-hip ratio

Participants with missing data: age n=1; sex n=1; Townsend Deprivation Index n=624; smoking n=1171; alcohol drinking n=1654; BMI n=10136; WC n=2161; HC n=2220; WHR n=2226; parental history of diabetes n=58299; fasting time n=1234.

#### Webtable 4. Associations of the first 11 metabolic biomarker principal components with risk of incident type 2 diabetes

| Principal component | Main model* HR (95% CI) | Main model + sequential adjustment † HR (95% CI) |
|---|---|---|
| 1 | 1.23 (1.17, 1.30) | 1.23 (1.17, 1.30) |
| 2 | 0.78 (0.73, 0.82) | 0.71 (0.67, 0.75) |
| 3 | 1.23 (1.18, 1.29) | 1.18 (1.13, 1.24) |
| 4 | 1.09 (1.04, 1.15) | 1.05 (1.00, 1.10) |
| 5 | 1.07 (1.02, 1.13) | 1.10 (1.04, 1.15) |
| 6 | 0.97 (0.92, 1.01) | 1.00 (0.95, 1.05) |
| 7 | 0.74 (0.70, 0.78) | 0.77 (0.73, 0.82) |
| 8 | 0.85 (0.81, 0.90) | 0.90 (0.86, 0.95) |
| 9 | 0.94 (0.90, 0.99) | 0.91 (0.87, 0.96) |
| 10 | 0.87 (0.83, 0.91) | 0.92 (0.88, 0.97) |
| 11 | 0.92 (0.87, 0.96) | 0.97 (0.93, 1.02) |

*Hazard ratios (95% CI) stratified by age-at-risk and sex and adjusted for assessment centre, Townsend deprivation index, smoking, alcohol drinking, body mass index, waist-to-hip ratio, fasting duration, and spectrometer.
† Hazard ratios (95% CI) stratified by age-at-risk and sex and adjusted for assessment centre, Townsend deprivation index, smoking, alcohol drinking, body mass index, waist-to-hip ratio, fasting duration, spectrometer, and preceding principal components. E.g. for principal component 3 the hazard ratio is stratified by age-at-risk and sex and adjusted for assessment centre, Townsend deprivation index, smoking, alcohol drinking, body mass index, waist-to-hip ratio, fasting duration, spectrometer, principal component 1 and principal component 2.

#### Webtable 5. Regression models for risk of incident type 2 diabetes

| | Basic model* | Basic model* plus metabolic biomarkers † | Extended model ‡ | Extended model ‡ plus metabolic biomarkers † |
|---|---|---|---|---|
| | HR (95% CI) | HR (95% CI) | HR (95% CI) | HR (95% CI) |
| Age (years) | | | | |
| <50 | 1.00 (reference) | 1.00 (reference) | 1.00 (reference) | 1.00 (reference) |
| 50 to <65 | 1.45 (1.28, 1.64) | 1.47 (1.30, 1.67) | 1.39 (1.23, 1.58) | 1.40 (1.23, 1.59) |
| ≥65 | 2.05 (1.76, 2.39) | 2.09 (1.79, 2.44) | 1.88 (1.61, 2.20) | 1.90 (1.62, 2.23) |
| Sex | | | | |
| Women | 1.00 (reference) | 1.00 (reference) | 1.00 (reference) | 1.00 (reference) |
| Men | 1.56 (1.42, 1.72) | 1.05 (0.94, 1.18) | 1.62 (1.47, 1.79) | 1.27 (1.12, 1.44) |
| Parental history of diabetes | | | | |
| No | 1.00 (reference) | 1.00 (reference) | 1.00 (reference) | 1.00 (reference) |
| Yes | 1.92 (1.73, 2.12) | 1.85 (1.67, 2.05) | 1.86 (1.68, 2.06) | 1.85 (1.67, 2.05) |
| Body mass index (kg/m²) | | | | |
| <25 | 1.00 (reference) | 1.00 (reference) | 1.00 (reference) | 1.00 (reference) |
| 25 to <30 | 2.45 (2.07, 2.90) | 1.83 (1.54, 2.18) | 1.64 (1.38, 1.96) | 1.52 (1.27, 1.82) |
| ≥30 | 6.71 (5.70, 7.90) | 3.89 (3.26, 4.64) | 2.69 (2.19, 3.30) | 2.32 (1.89, 2.86) |
| HbA1c (%) | | | | |
| <6.0 (normal) | 1.00 (reference) | 1.00 (reference) | 1.00 (reference) | 1.00 (reference) |
| ≥6.0 (pre-diabetes) | 10.09 (9.02, 11.29) | 8.61 (7.68, 9.64) | 8.80 (7.86, 9.85) | 8.25 (7.37, 9.24) |
| Principal components | | | | |
| 1 | | 1.26 (1.19, 1.34) | | 1.16 (1.08, 1.25) |
| 2 | | 0.71 (0.67, 0.76) | | 0.82 (0.76, 0.88) |
| 3 | | 1.12 (1.07, 1.17) | | 1.11 (1.06, 1.17) |
| 4 | | 1.01 (0.96, 1.06) | | 1.00 (0.95, 1.05) |
| 5 | | 1.08 (1.03, 1.14) | | 1.07 (1.02, 1.12) |
| 6 | | 1.01 (0.96, 1.05) | | 1.00 (0.96, 1.05) |
| 7 | | 0.76 (0.73, 0.80) | | 0.80 (0.76, 0.84) |
| 8 | | 0.92 (0.87, 0.97) | | 0.92 (0.88, 0.97) |
| 9 | | 0.92 (0.88, 0.97) | | 0.91 (0.87, 0.96) |
| 10 | | 0.92 (0.88, 0.97) | | 0.93 (0.89, 0.98) |
| 11 | | 1.01 (0.97, 1.06) | | 1.02 (0.98, 1.07) |
| Blood pressure | | | | |
| SBP ≤130 mmHg and DBP ≤85 mmHg and no antihypertensive medications | | | 1.00 (reference) | 1.00 (reference) |
| SBP >130 mmHg or DBP >85 mmHg or on antihypertensive medication | | | 1.48 (1.30, 1.69) | 1.42 (1.25, 1.62) |
| Waist circumference (cm) | | | | |
| ≤102 in men; ≤88 in women | | | 1.00 (reference) | 1.00 (reference) |
| >102 in men; >88 in women | | | 1.82 (1.59, 2.08) | 1.70 (1.49, 1.95) |
| HDL-cholesterol (mmol/L) | | | | |
| ≥1.0344 in men; ≥1.293 in women | | | 1.00 (reference) | 1.00 (reference) |
| <1.0344 in men; <1.293 in women | | | 1.80 (1.62, 1.99) | 1.38 (1.21, 1.58) |
| Triglycerides (mmol/L) | | | | |
| <1.6935 | | | 1.00 (reference) | 1.00 (reference) |
| ≥1.6935 | | | 1.57 (1.41, 1.75) | 1.12 (0.96, 1.31) |

*Basic model: Hazard ratios (95% CI) adjusted for age, sex, parental history of diabetes, BMI and HbA1c
† Metabolic biomarkers comprise the first 11 metabolic biomarker principal components
‡ Extended model: Hazard ratios (95% CI) adjusted for age, sex, parental history of diabetes, BMI, HbA1c, waist circumference, blood pressure, triglycerides, HDL-cholesterol
DBP= diastolic blood pressure; SBP= systolic blood pressure

#### Webtable 6. Performance of risk prediction models for incident type 2 diabetes among participants taking lipid-lowering medication at recruitment

| Performance metric | Basic model* | Basic model* plus metabolic biomarkers † | Extended model ‡ | Extended model ‡ plus metabolic biomarkers † |
|---|---|---|---|---|
| **C-statistic (CI) §** | 0.761 (0.747, 0.777) | 0.789 (0.779, 0.806) | 0.782 (0.768, 0.798) | 0.793 (0.783, 0.810) |
| **Metrics of relative performance** | | | | |
| $\chi^2$ \|\| # | 174 (p<0.0001) | | 88 (p<0.0001) | |
| %increase $\chi^2$ | 15 | | 7 | |
| Absolute IDI # $ | 1.3 (0.6, 2.0) | | 0.7 (0.3, 1.1) | |
| Relative IDI (%) (CI) # $ | 9.1 (4.7, 14.2) | | 4.4 (1.8, 8.0) | |
| Continuous NRI (CI) # ** | | | | |
| Events | 0.13 (0.10, 0.21) | | 0.07 (0.03, 0.15) | |
| Non-events | 0.24 (0.20, 0.29) | | 0.13 (0.08, 0.17) | |
| Overall | 0.38 (0.33, 0.48) | | 0.21 (0.11, 0.29) | |

* Basic model: age, sex, parental history of diabetes, body mass index, HbA1c
† Metabolic biomarkers comprise the first 11 metabolic biomarker principal components
‡ Extended model: basic model plus waist circumference, blood pressure, triglycerides, HDL-cholesterol
§ The **c-statistic** measures the ability of a model to rank participants from low to high risk. Given two randomly selected individuals, one who develops T2D and one who does not, the c-statistic is the probability that the model will give a higher predicted risk for the individual who develops T2D. An uninformative model will have a c-statistic of 0.5 and a model that discriminates perfectly will have a c-statistic of 1.0.
|| 11 DF
### Bias-corrected estimates and confidence intervals were derived using 200 bootstrap samples
$ The **IDI** quantifies the difference between two models in their ability to predict risk. It is calculated as the difference between the two models in the mean predicted T2D risk among those who did develop T2D minus the mean predicted risk of T2D in those who did not develop T2D (i.e., it is the difference between two differences). When metabolic biomarkers were added to the basic model, the separation in mean predicted T2D risk between those who did develop T2D, compared with those who did not develop T2D, increased in relative terms by 9.1%. Positive IDI values indicate improved T2D risk classification following addition of metabolic biomarkers to the risk prediction model.
** The continuous **NRI** quantifies the appropriateness of the change in predicted probabilities of T2D between two models. The ‘Events’ NRI is calculated among those who developed T2D, and the ‘Non-events’ NRI is calculated among those who did not develop T2D. Both statistics are calculated as the probability of an ‘appropriate’ change in predicted risk (after addition of metabolic biomarkers to the model) minus the probability of an ‘inappropriate’ change in predicted risk. For those who developed T2D, an appropriate change would be a higher predicted T2D risk after addition of metabolic biomarkers to the model. An inappropriate change would be a lower predicted T2D risk after addition of metabolic biomarkers to the model. When metabolic biomarkers were added to the basic model, among those w ho developed T2D, 13% more were assigned a higher predicted T2D risk than were assigned a lower predicted risk. The overall NRI is th e sum of the ‘Events’ and ‘Non-events’ NRI statistics. Positive NRI values indicate that addition of metabolic biomarkers results in a superior model.
DF= degrees of freedom; IDI= integrated discrimination improvement; NRI= net reclassification improvement; T2D= type 2 diabetes

#### Webfigure 1. Participant exclusions to derive analysis population

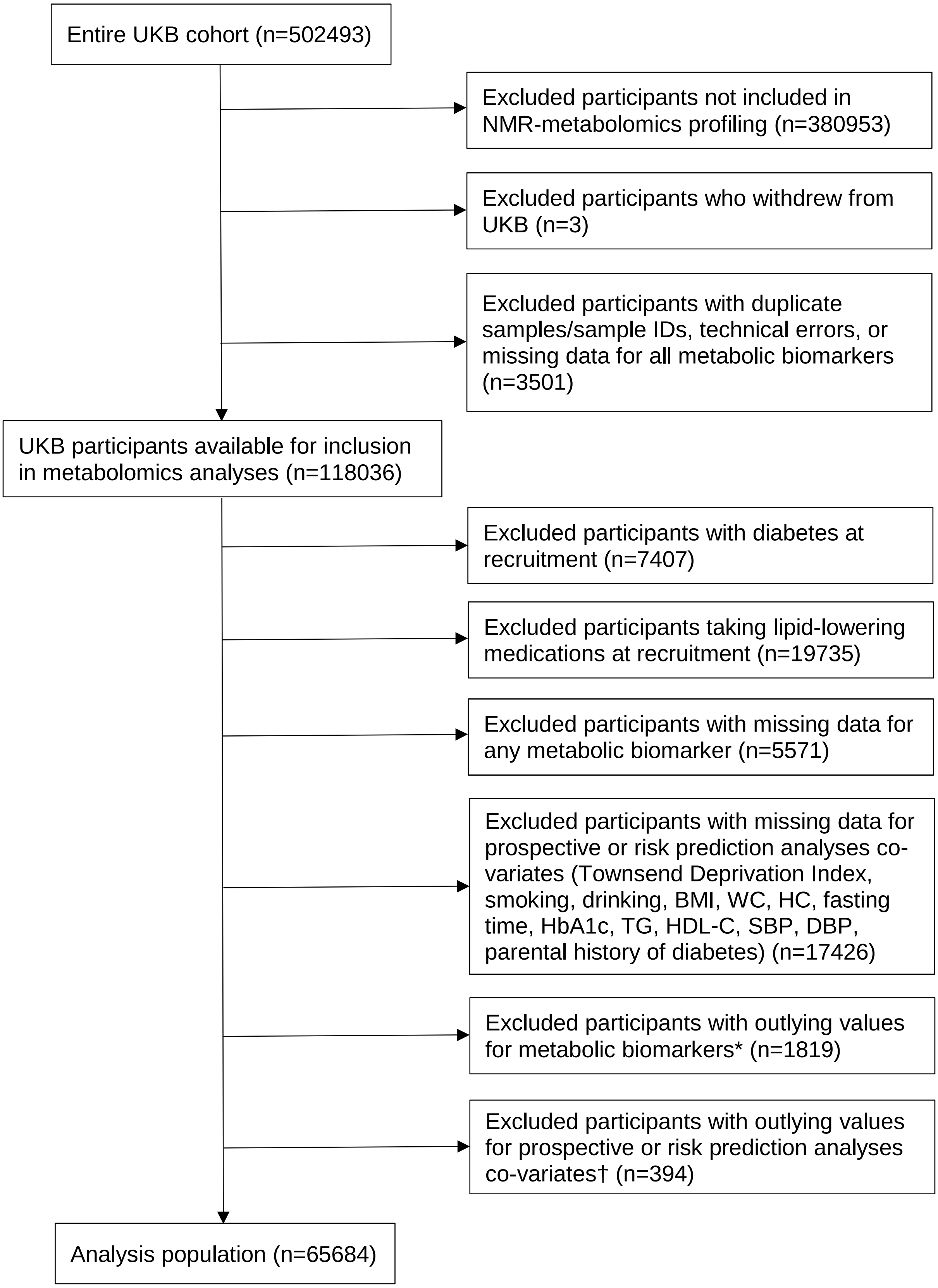


* Values >4 standard deviations from the mean and within the top or bottom 0.003% of the original or log-transformed metabolic biomarker distribution

† Weight <40 or ≥150 kg; height <140 or ≥200 cm; BMI <15 or ≥50 kg/m2; WC <55 or ≥145 cm; HC <75 or ≥150 cm; WHR <0.60 or ≥1.15

BMI=body mass index; DBP=diastolic blood pressure; HC=hip circumference; HDL-C=HDL-cholesterol from routine clinical chemistry measures; SBP=systolic blood pressure; TG=triglycerides from routine clinical chemistry measures; UKB=UK Biobank; WC=waist circumference

### Webfigure 2. Cross-correlations of metabolic biomarkers

#### a) Lipoproteins, fatty acids and other lipids, and small molecules

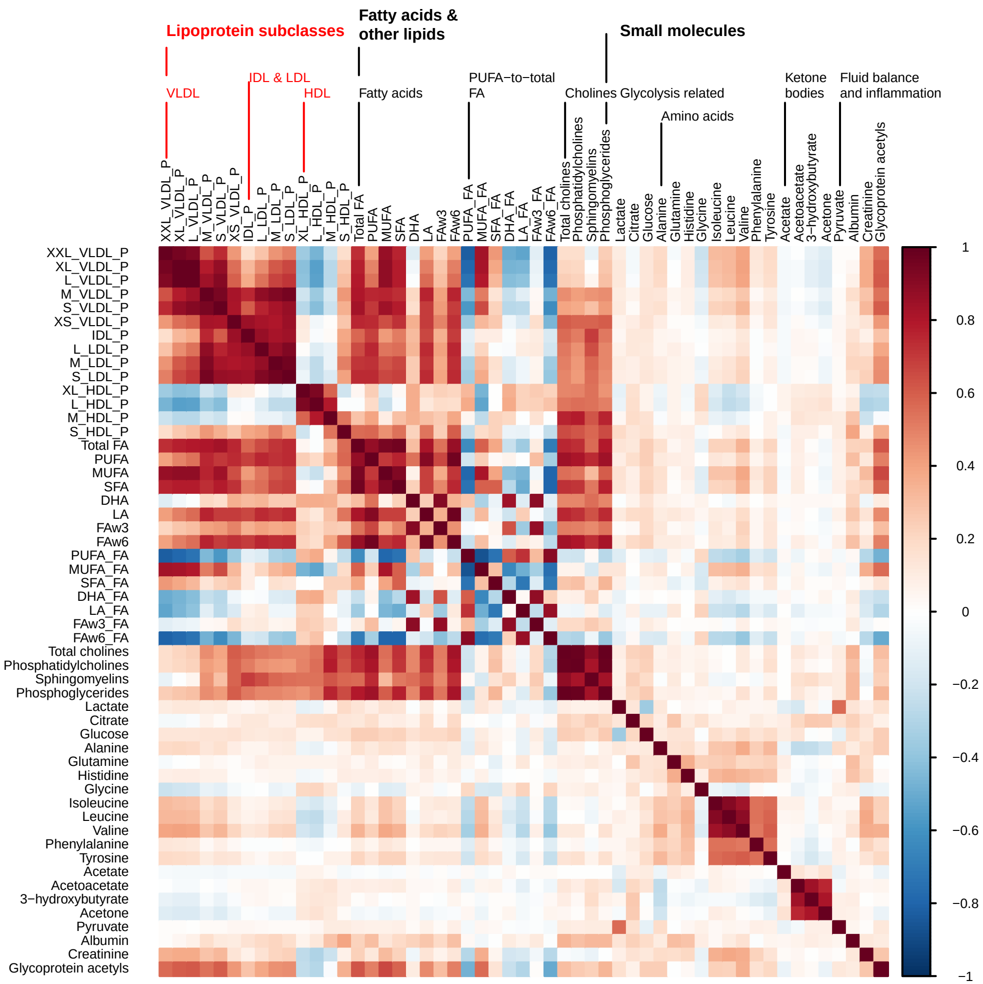


#### b) Lipoproteins, lipids by lipoprotein subclasses, particle sizes, and apolipoproteins

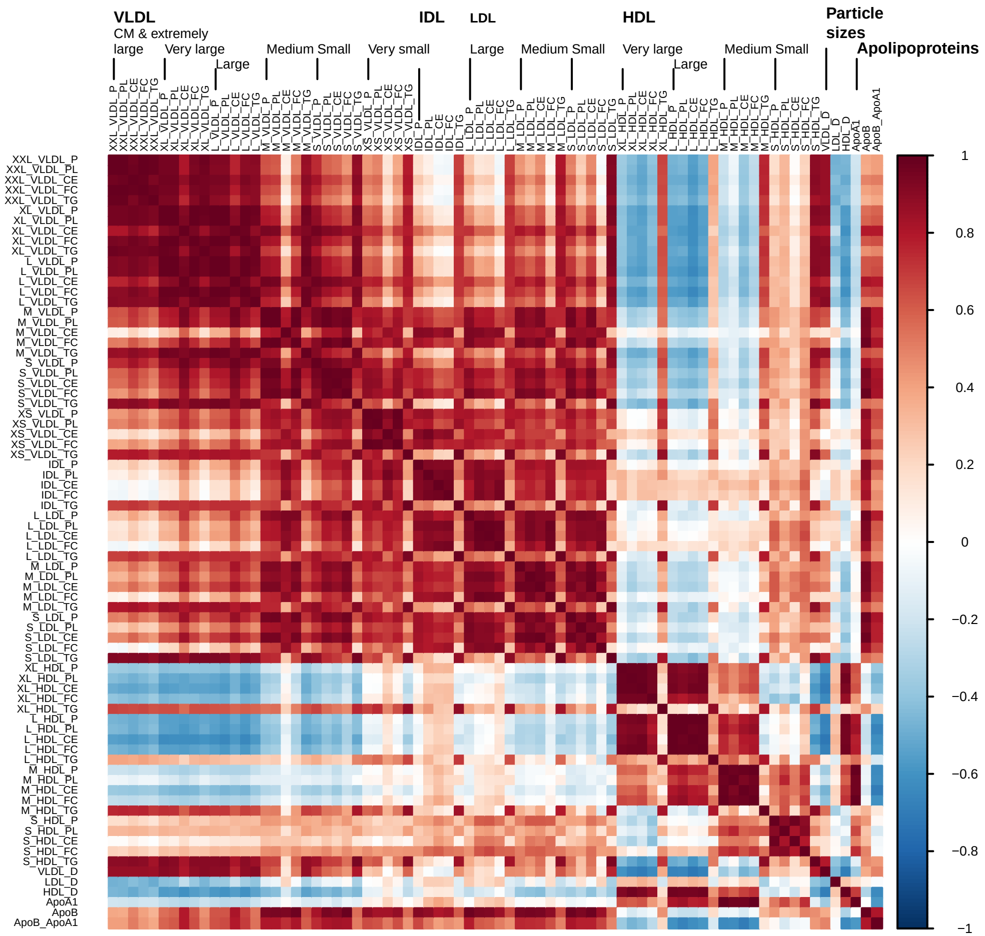


Apo−A1=apolipoprotein A1; Apo−B=apolipoprotein B; DHA=docosahexaenoic acid; FA=fatty acids; FAw3=omega−3 fatty acids; FAw6=omega−6 fatty acids; HDL=high density lipoproteins; HDL−D=high density lipoprotein particle diameter; IDL=intermediate density lipoproteins; L=large; LA=linoleic acid; LDL=low density lipoproteins; LDL−D=low density lipoprotein particle diameter; LP=lipoprotein; M=medium; MUFA=monounsaturated fatty acids; PUFA=polyunsaturated fatty acids; S=small; SFA=saturated fatty acids; T2D=type 2 diabetes; VLDL=very low density lipoproteins; VLDL−D=very low density lipoprotein particle diameter; XL=very large; XS=very small; XXL=extremely large

### Webfigure 3. Associations of metabolic biomarkers with risk of incident type 2 diabetes

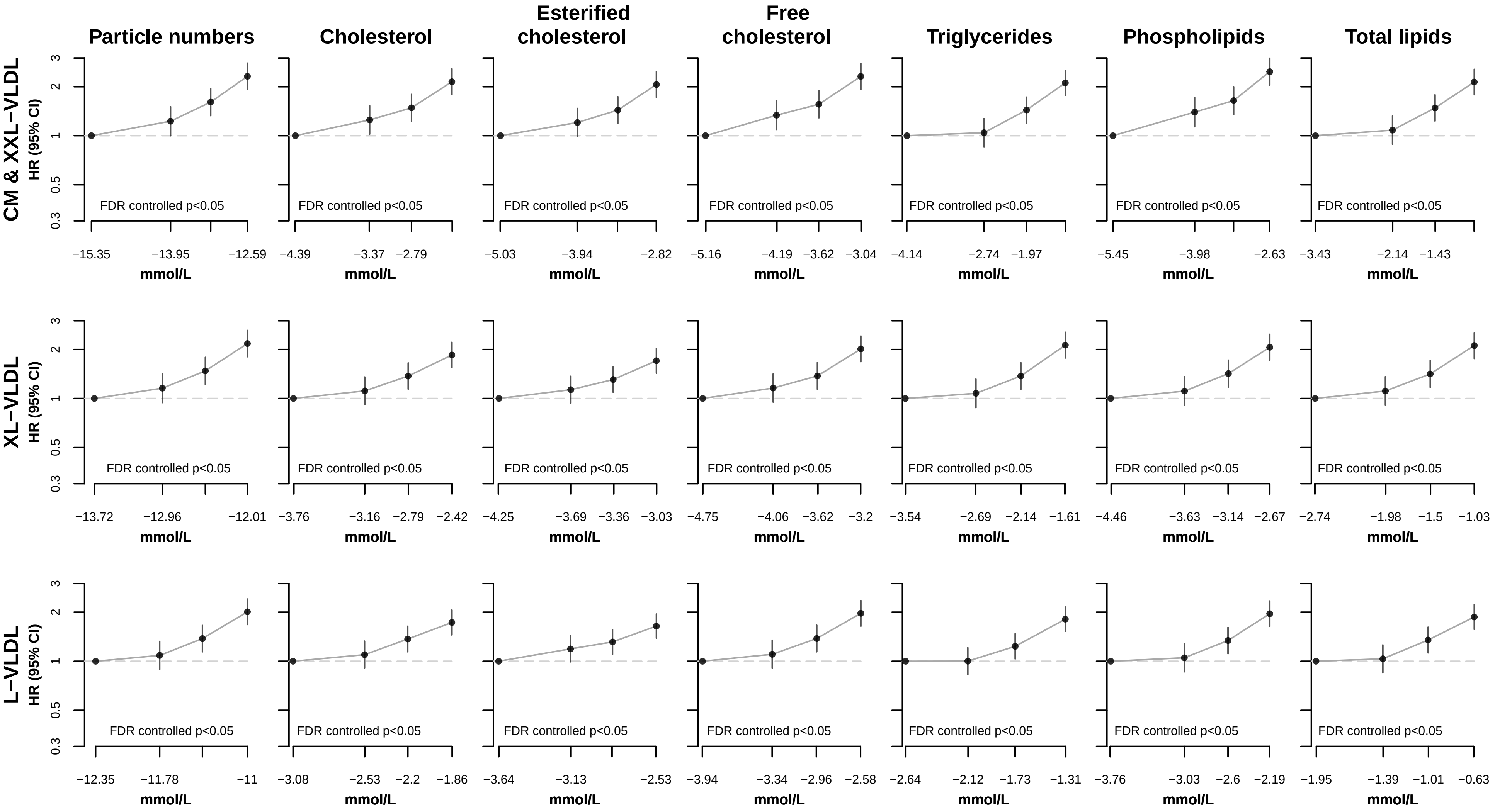


Hazard ratios (HR) stratified by age−at−risk and sex and adjusted for assessment centre, Townsend deprivation index, smoking ,alcohol drinking, body mass index, waist−to−hip ratio, fasting duration, and spectorometer.
Numbers on the x−axis correspond to median values within each quartile for each metabolic biomarker on the natural log scale.
Circles represent the HR and vertical lines indicate the 95% CI.
CM= chylomicrons; FA= fatty acids; HDL= high density lipoproteins; IDL= intermediate density lipoproteins; L= large; LDL= low density liporpoteins; M= medium; S= small; VLDL= very low density lipoproteins; XL= very large; XS= very small; XXL= extremely large

### Webfigure 3. Associations of metabolic biomarkers with risk of incident type 2 diabetes

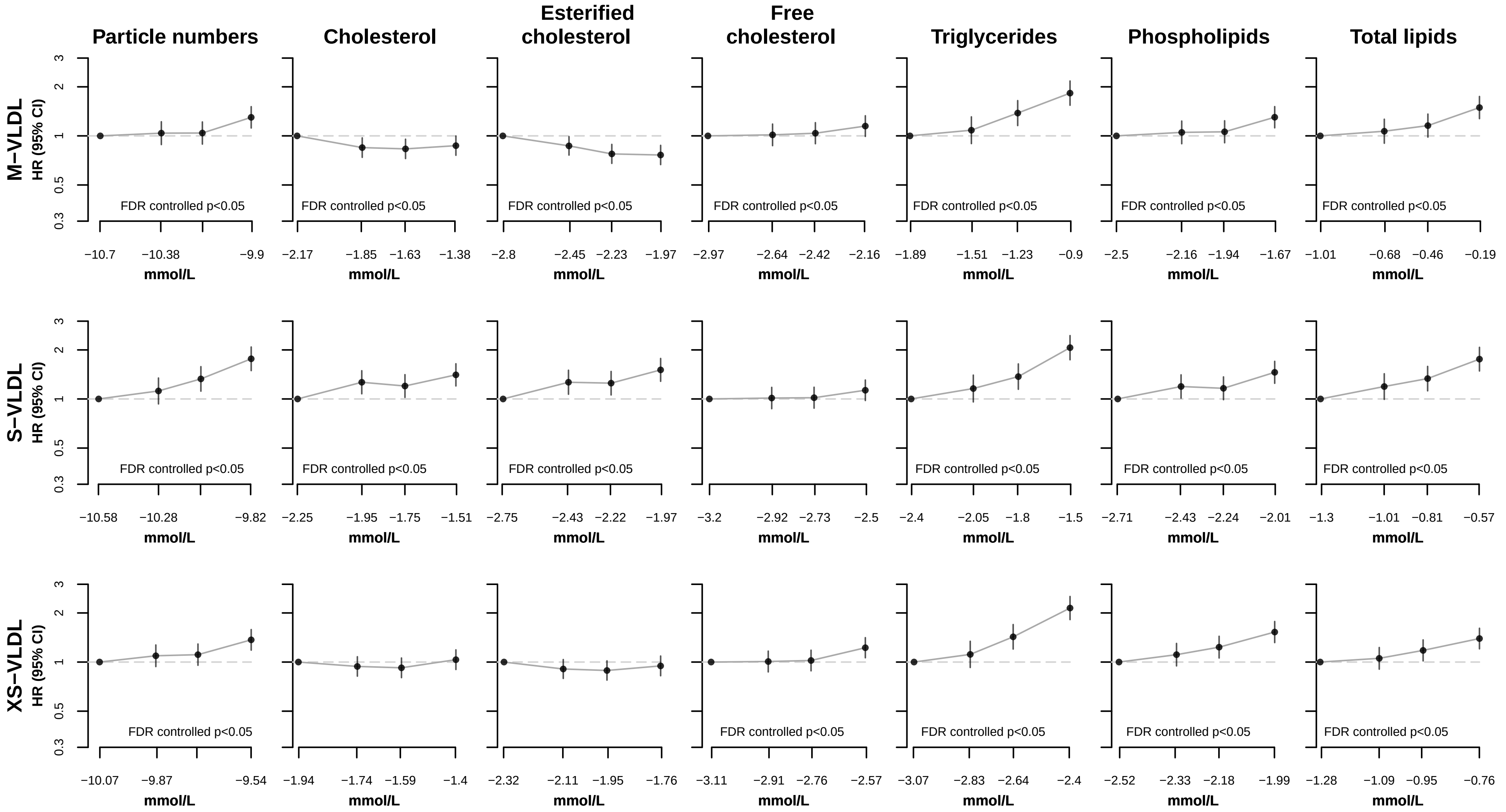


Hazard ratios (HR) stratified by age−at−risk and sex and adjusted for assessment centre, Townsend deprivation index, smoking ,alcohol drinking, body mass index, waist−to−hip ratio, fasting duration, and spectorometer.
Numbers on the x−axis correspond to median values within each quartile for each metabolic biomarker on the natural log scale.
Circles represent the HR and vertical lines indicate the 95% CI.
CM= chylomicrons; FA= fatty acids; HDL= high density lipoproteins; IDL= intermediate density lipoproteins; L= large; LDL= low density liporpoteins; M= medium; S= small; VLDL= very low density lipoproteins; XL= very large; XS= very small; XXL= extremely large

### Webfigure 3. Associations of metabolic biomarkers with risk of incident type 2 diabetes

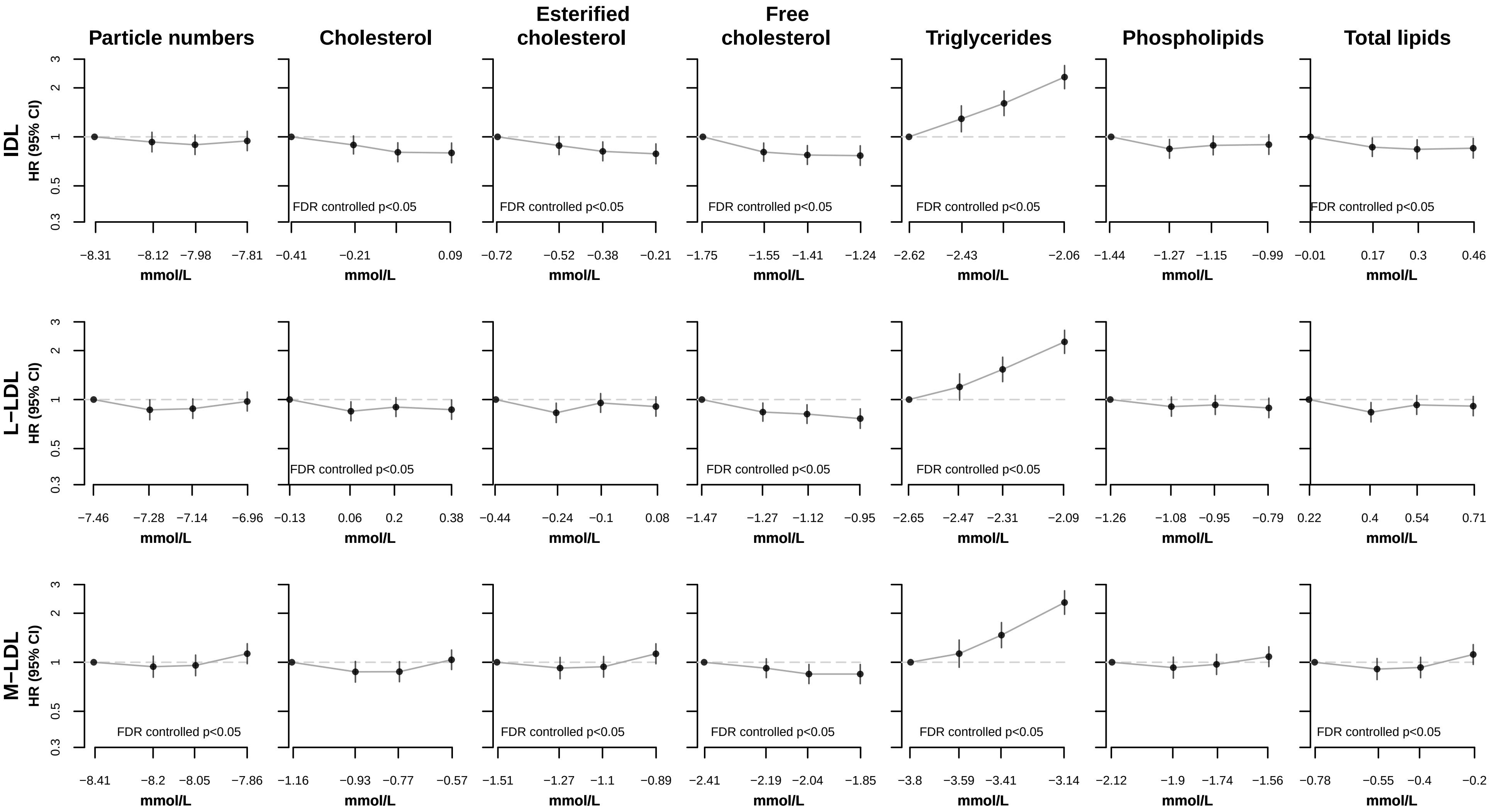


Hazard ratios (HR) stratified by age−at−risk and sex and adjusted for assessment centre, Townsend deprivation index, smoking ,alcohol drinking, body mass index, waist−to−hip ratio, fasting duration, and spectorometer.
Numbers on the x−axis correspond to median values within each quartile for each metabolic biomarker on the natural log scale.
Circles represent the HR and vertical lines indicate the 95% CI.
CM= chylomicrons; FA= fatty acids; HDL= high density lipoproteins; IDL= intermediate density lipoproteins; L= large; LDL= low density liporpoteins; M= medium; S= small; VLDL= very low density lipoproteins; XL= very large; XS= very small; XXL= extremely large

### Webfigure 3. Associations of metabolic biomarkers with risk of incident type 2 diabetes

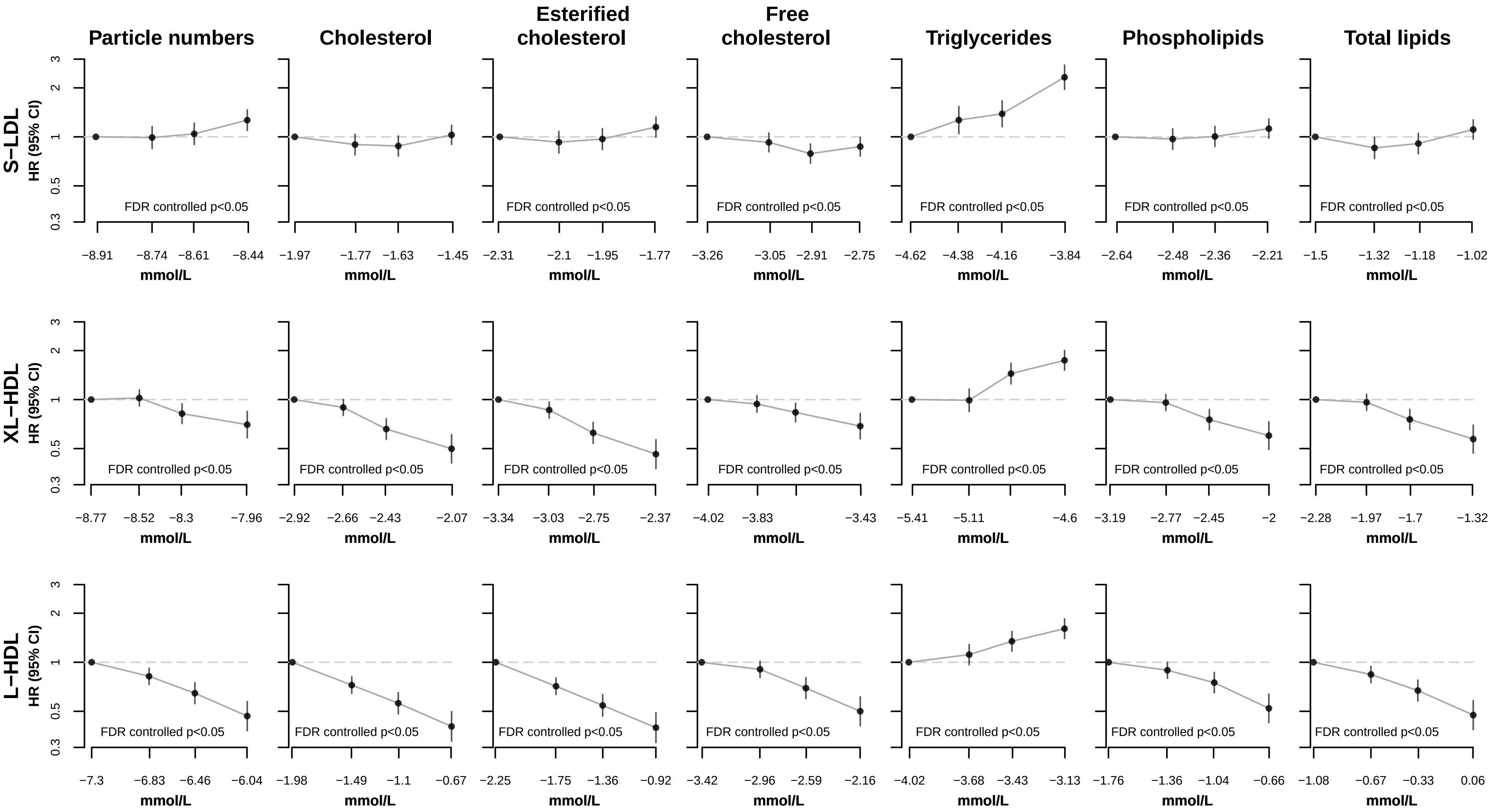


Hazard ratios (HR) stratified by age−at−risk and sex and adjusted for assessment centre, Townsend deprivation index, smoking ,alcohol drinking, body mass index, waist−to−hip ratio, fasting duration, and spectorometer.
Numbers on the x−axis correspond to median values within each quartile for each metabolic biomarker on the natural log scale.
Circles represent the HR and vertical lines indicate the 95% CI.
CM= chylomicrons; FA= fatty acids; HDL= high density lipoproteins; IDL= intermediate density lipoproteins; L= large; LDL= low density liporpoteins; M= medium; S= small; VLDL= very low density lipoproteins; XL= very large; XS= very small; XXL= extremely large

### Webfigure 3. Associations of metabolic biomarkers with risk of incident type 2 diabetes

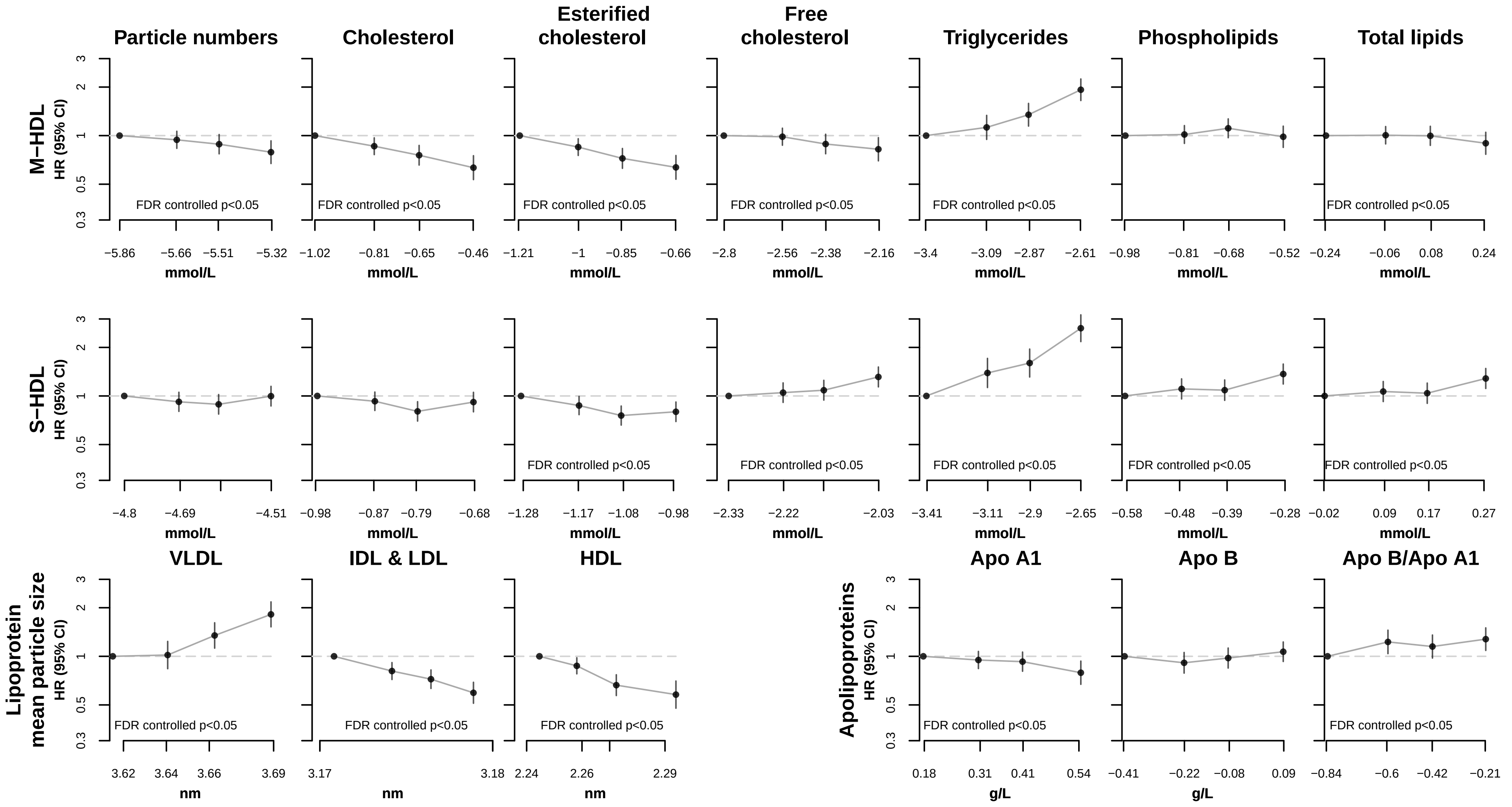


Hazard ratios (HR) stratified by age−at−risk and sex and adjusted for assessment centre, Townsend deprivation index, smoking ,alcohol drinking, body mass index, waist−to−hip ratio, fasting duration, and spectorometer.
Numbers on the x−axis correspond to median values within each quartile for each metabolic biomarker on the natural log scale.
Circles represent the HR and vertical lines indicate the 95% CI.
CM= chylomicrons; FA= fatty acids; HDL= high density lipoproteins; IDL= intermediate density lipoproteins; L= large; LDL= low density liporpoteins; M= medium; S= small; VLDL= very low density lipoproteins; XL= very large; XS= very small; XXL= extremely large

### Webfigure 3. Associations of metabolic biomarkers with risk of incident type 2 diabetes

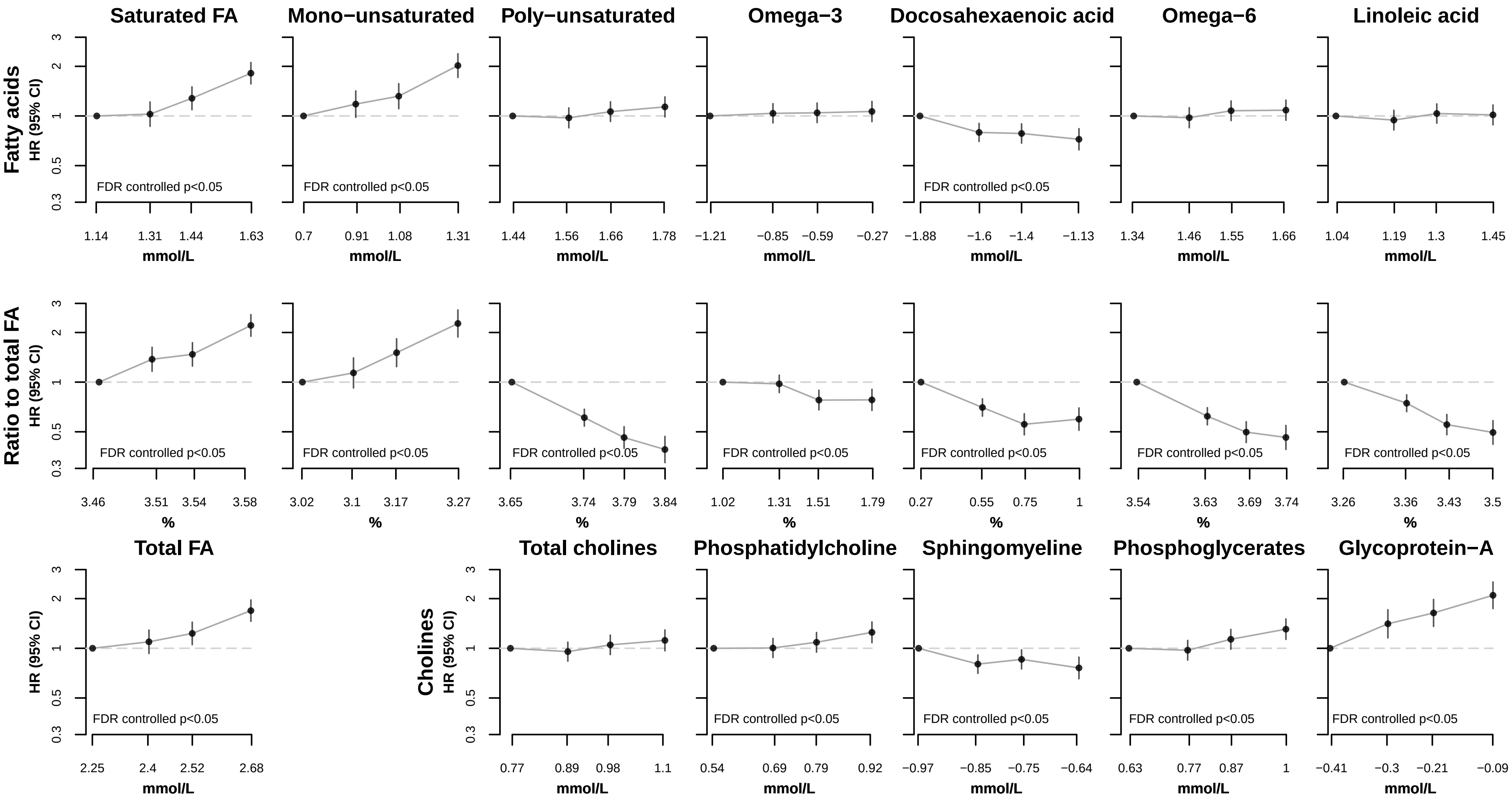


Hazard ratios (HR) stratified by age−at−risk and sex and adjusted for assessment centre, Townsend deprivation index, smoking ,alcohol drinking, body mass index, waist−to−hip ratio, fasting duration, and spectorometer.
Numbers on the x−axis correspond to median values within each quartile for each metabolic biomarker on the natural log scale.
Circles represent the HR and vertical lines indicate the 95% CI.
CM= chylomicrons; FA= fatty acids; HDL= high density lipoproteins; IDL= intermediate density lipoproteins; L= large; LDL= low density liporpoteins; M= medium; S= small; VLDL= very low density lipoproteins; XL= very large; XS= very small; XXL= extremely large

### Webfigure 3. Associations of metabolic biomarkers with risk of incident type 2 diabetes

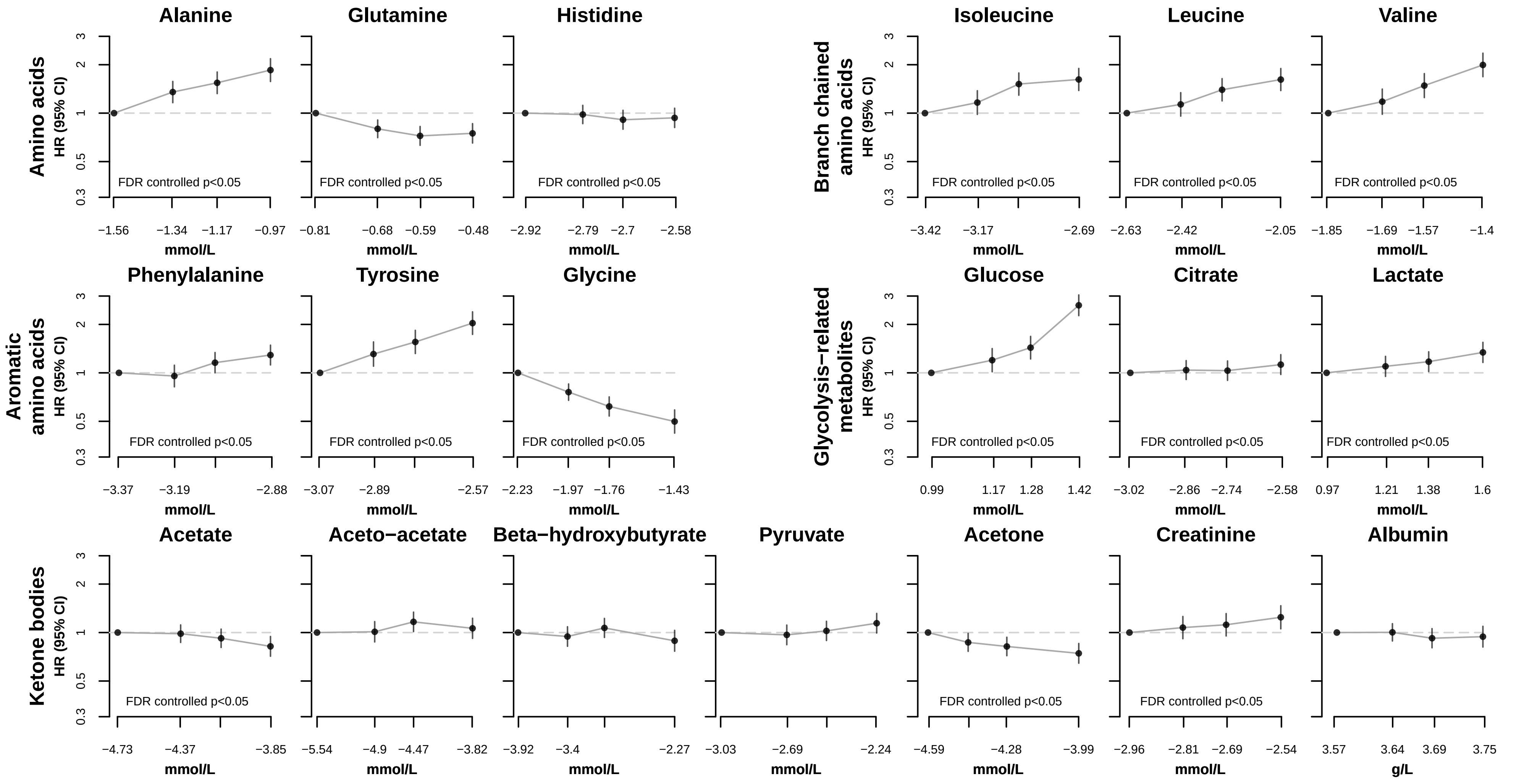


Hazard ratios (HR) stratified by age−at−risk and sex and adjusted for assessment centre, Townsend deprivation index, smoking ,alcohol drinking, body mass index, waist−to−hip ratio, fasting duration, and spectorometer.
Numbers on the x−axis correspond to median values within each quartile for each metabolic biomarker on the natural log scale.
Circles represent the HR and vertical lines indicate the 95% CI.
CM= chylomicrons; FA= fatty acids; HDL= high density lipoproteins; IDL= intermediate density lipoproteins; L= large; LDL= low density liporpoteins; M= medium; S= small; VLDL= very low density lipoproteins; XL= very large; XS= very small; XXL= extremely large

### Webfigure 4. Associations of metabolic biomarkers with risk of incident type 2 diabetes by sex

#### a) HR of incident T2D per 1-SD higher metabolic biomarker on the natural log scale

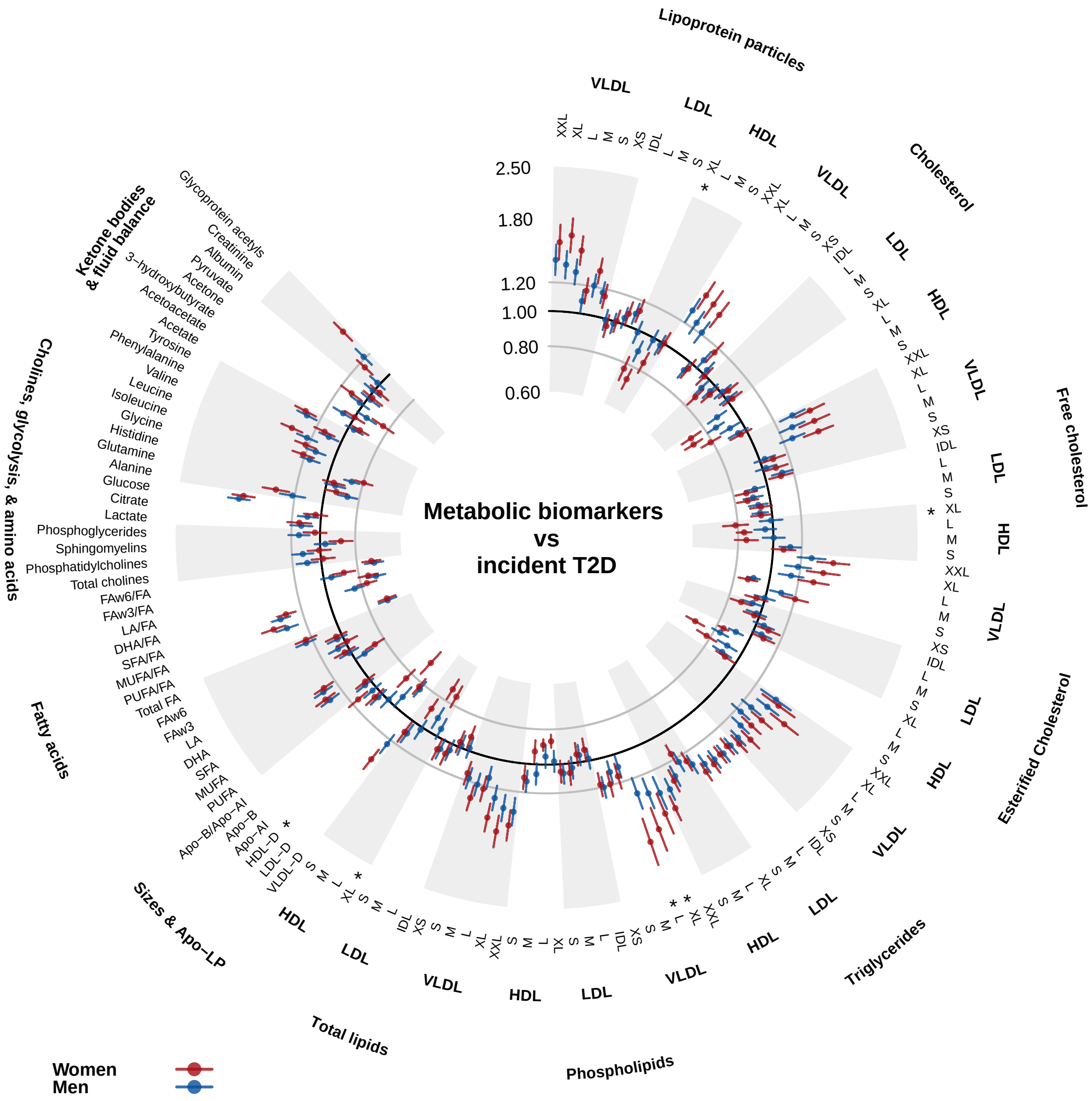


#### b) Effect size estimates for incident T2D per 1-SD higher metabolic biomarker on the natural log scale

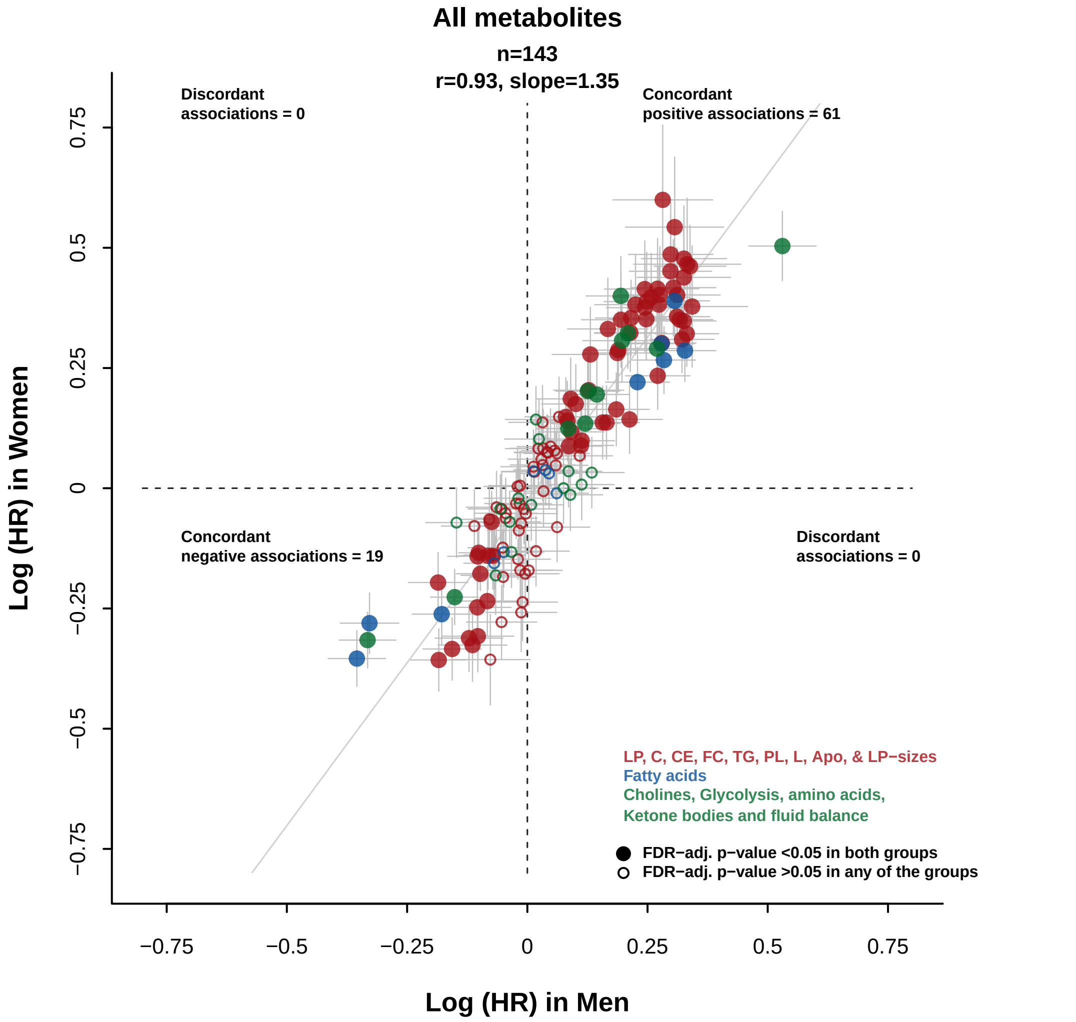


Hazard ratios (HR) stratified by age−at−risk and adjusted for assessment centre, Townsend deprivation index, smoking, alcohol drinking, body mass index, waist−to−hip ratio, fasting duration and spectrometer

* p for heterogeneity <0.05 after controlling for false discovery rate

911 men and 808 women developed incident T2D

Apo=apolipoproteins; Apo−A1=apolipoprotein A1; Apo−B=apolipoprotein B; C=cholesterol; CE=esterified cholesterol; DHA=docosahexaenoic acid; FA=fatty acids; FAw3=omega−3 fatty acids; FAw6=omega−6 fatty acids; FC=free cholesterol; FDR=false discovery rate; HDL=high density lipoproteins; HDL−D=high density lipoprotein particle diameter; IDL=intermediate density lipoproteins; L=large; LA=linoleic acid; LDL=low density lipoproteins; LDL−D=low density lipoprotein particle diameter; LP=lipoprotein; M=medium; MUFA=monounsaturated fatty acids; PUFA=polyunsaturated fatty acids; S=small; SFA=saturated fatty acids; T2D=type 2 diabetes; TG=triglycerides; VLDL=very low density lipoproteins; VLDL−D=very low density lipoprotein particle diameter; XL=very large; XS=very small; XXL=extremely large

#### Webfigure 5. Associations of metabolic biomarkers with risk of incident type 2 diabetes by age

**a) HR of incident T2D per 1-SD higher metabolic biomarker on the natural log scale**

**b) Effect size estimates for incident T2D per 1-SD higher metabolic biomarker on the natural log scale**

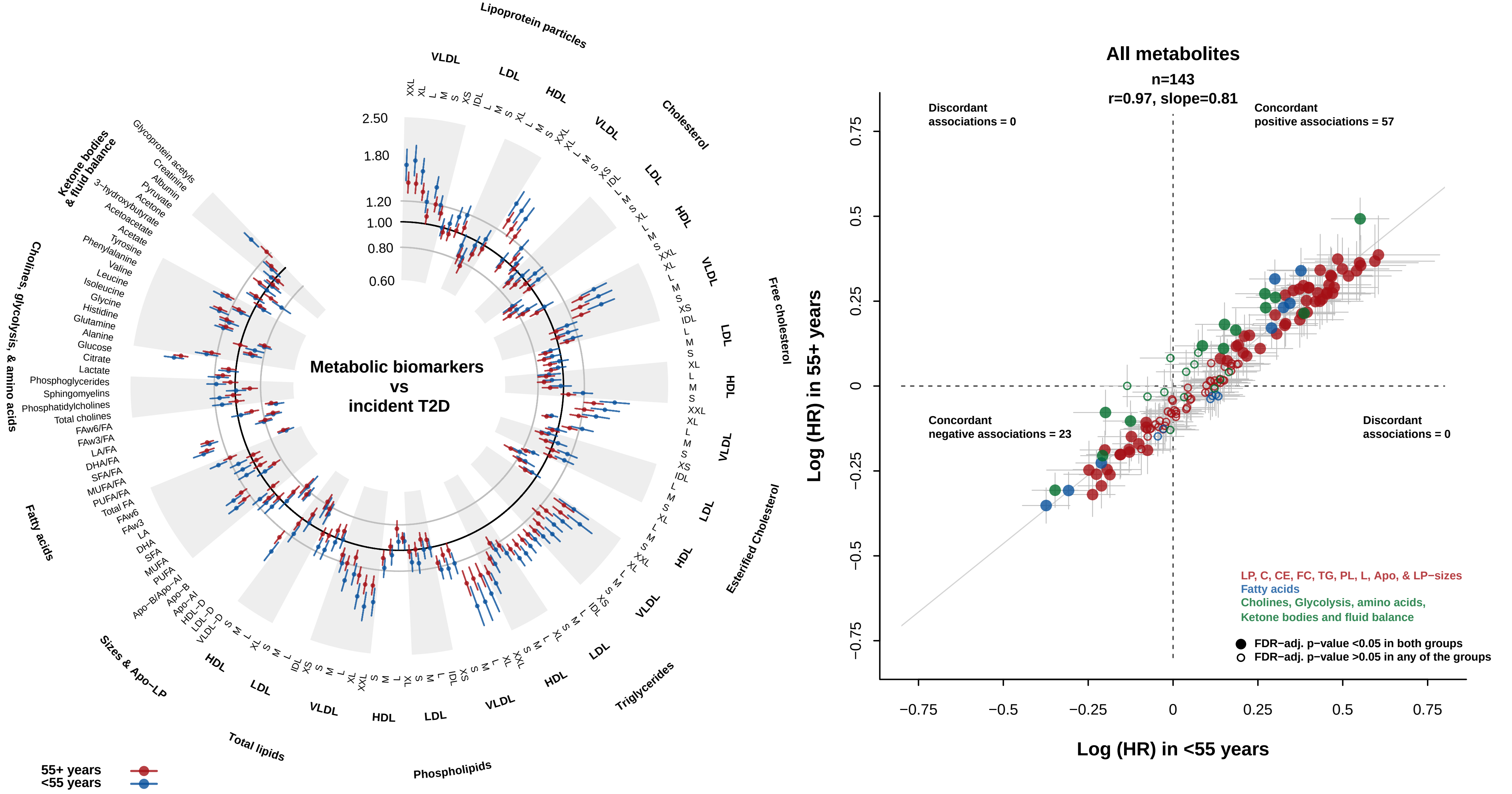


Hazard ratios (HR) stratified by age−at−risk and adjusted for assessment centre, Townsend deprivation index, smoking, alcohol drinking, body mass index, waist−to−hip ratio, fasting duration and spectrometer

* p for heterogeneity <0.05 after controlling for false discovery rate

616 participants <55 years at baseline and 1103 participants 55+ years at baseline developed incident T2D

Apo=apolipoproteins; Apo−A1=apolipoprotein A1; Apo−B=apolipoprotein B; C=cholesterol; CE=esterified cholesterol; DHA=docosahexaenoic acid; FA=fatty acids; FAw3=omega−3 fatty acids; FAw6=omega−6 fatty acids; FC=free cholesterol; FDR=false discovery rate; HDL=high density lipoproteins; HDL−D=high density lipoprotein particle diameter; IDL=intermediate density lipoproteins; L=large; LA=linoleic acid; LDL=low density lipoproteins; LDL−D=low density lipoprotein particle diameter; LP=lipoprotein; M=medium; MUFA=monounsaturated fatty acids; PUFA=polyunsaturated fatty acids; S=small; SFA=saturated fatty acids; T2D=type 2 diabetes; TG=triglycerides; VLDL=very low density lipoproteins; VLDL−D=very low density lipoprotein particle diameter; XL=very large; XS=very small; XXL=extremely large

#### Webfigure 6. Associations of metabolic biomarkers with risk of incident type 2 diabetes excluding the first three years of follow−up

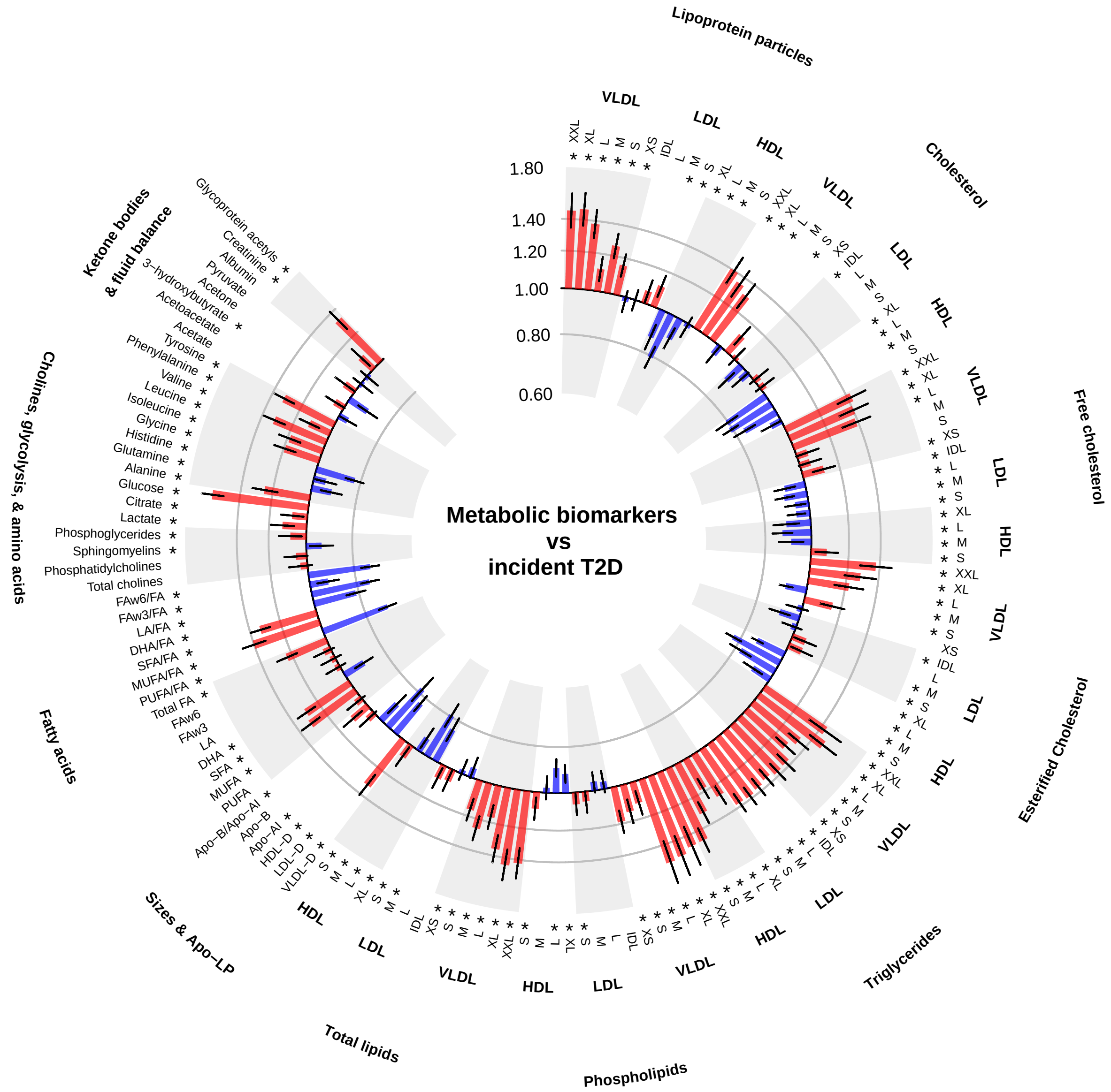


Hazard ratios are presented per 1−SD higher metabolic biomarkers on the natural log scale, stratified by age−at−risk and sex and adjusted for assessment centre, Townsend deprivation index, smoking, alcohol drinking, body mass index, waist−to−hip ratio, fasting duration, and spectrometer

*false discovery rate controlled p<0.05

Apo−A1=apolipoprotein A1; Apo−B=apolipoprotein B; DHA=docosahexaenoic acid; FA=fatty acids; FAw3=omega−3 fatty acids; FAw6=omega−6 fatty acids; HDL=high density lipoproteins; HDL−D=high density lipoprotein particle diameter; IDL=intermediate density lipoproteins; L=large; LA=linoleic acid; LDL=low density lipoproteins; LDL−D=low density lipoprotein particle diameter; LP=lipoprotein; M=medium; MUFA=monounsaturated fatty acids; PUFA=polyunsaturated fatty acids; S=small; SFA=saturated fatty acids; T2D=type 2 diabetes; VLDL=very low density lipoproteins; VLDL−D=very low density lipoprotein particle diameter; XL=very large; XS=very small; XXL=extremely large

#### Webfigure 7. Importance of the first 20 metabolic biomarker principal components

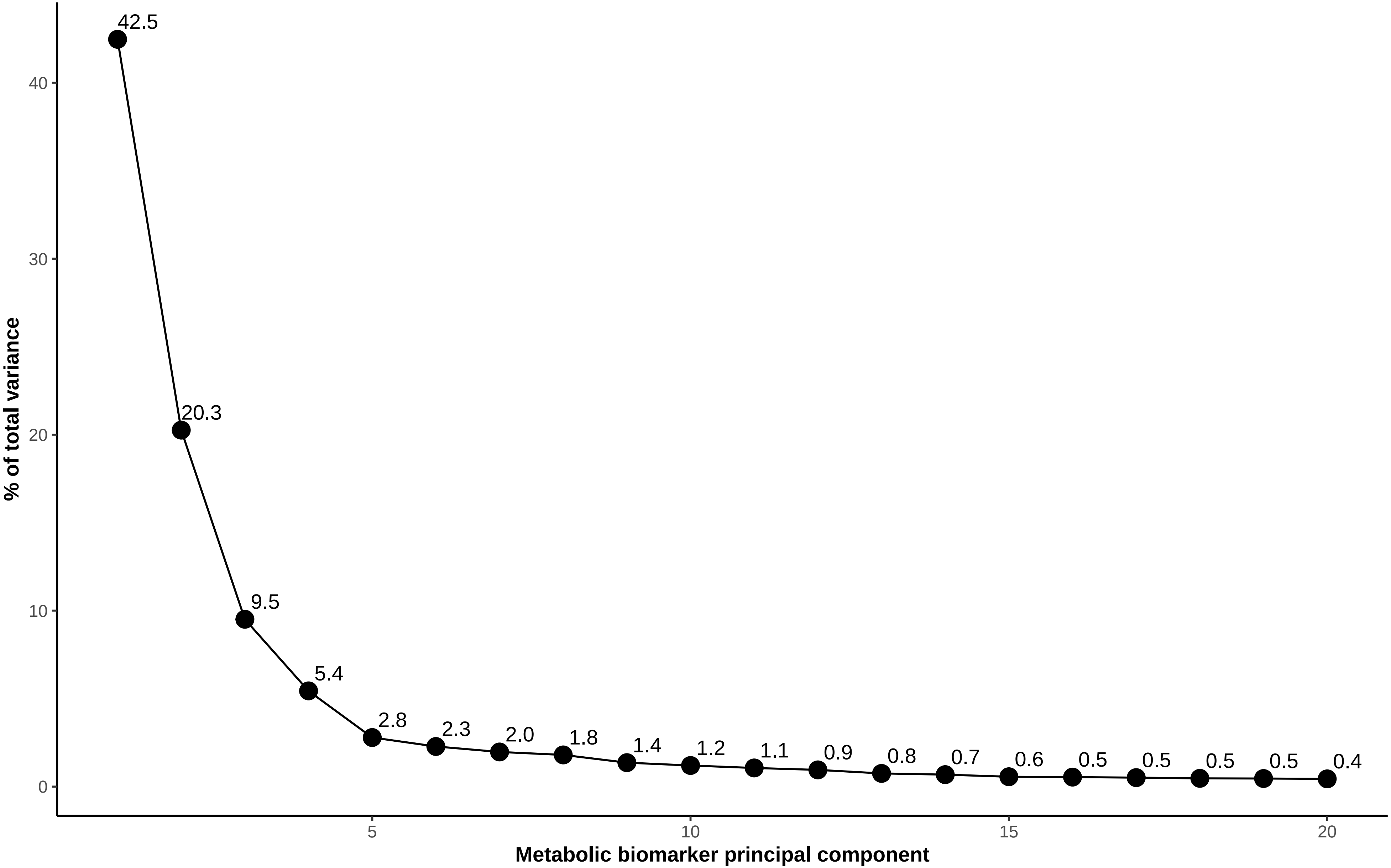


Numbers adjacent to the points are the percentages of total variance in individual biomarkers explained by each principal component.

### Webfigure 8. Characterisation of the first 11 metabolic biomarker principal components

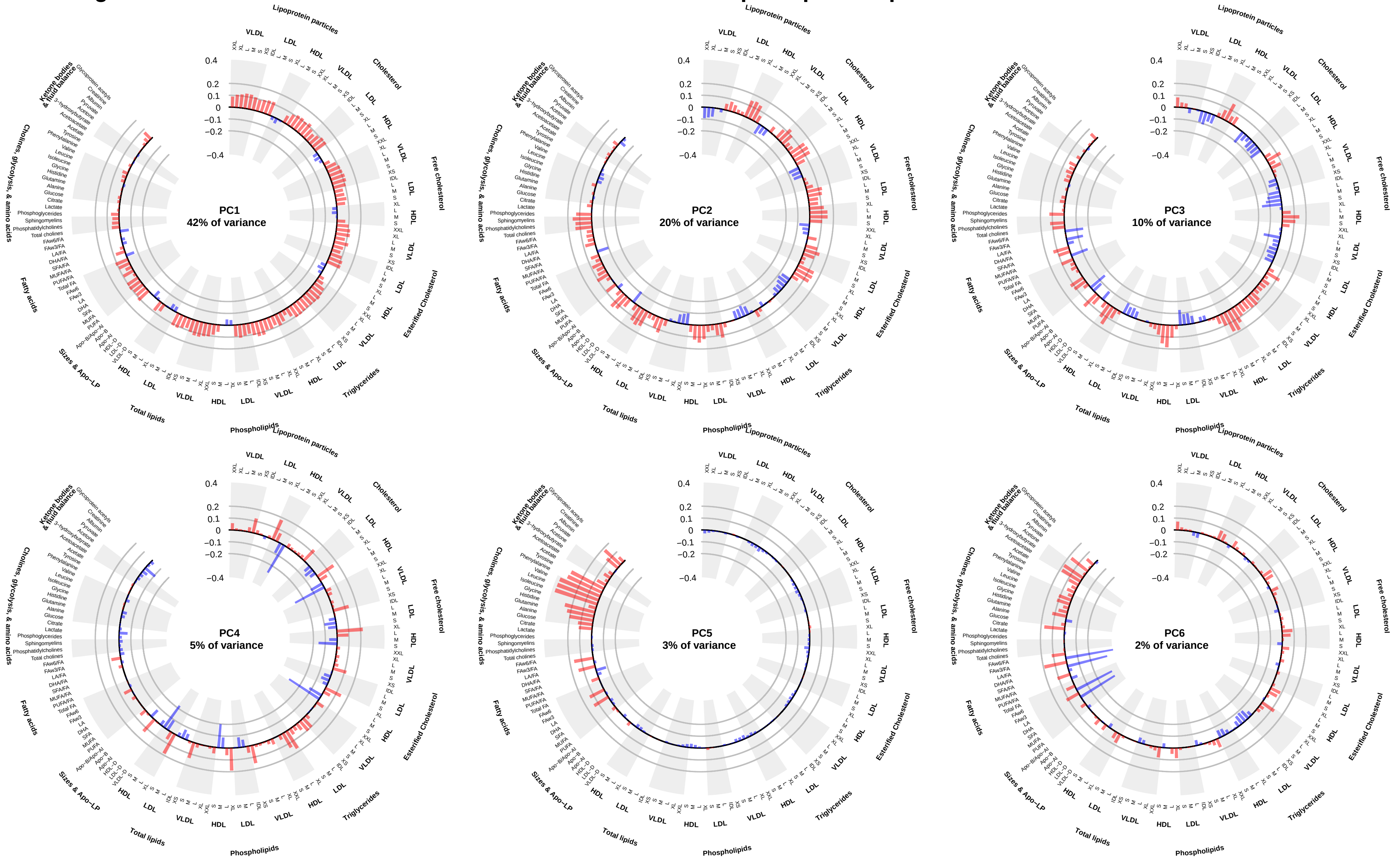


Factor loadings for each principal component are presented, representing the coefficients of the metabolic biomarkers from which each principal component was calculated. The first 6 principal components explain 83 % of the variance. Apo−A1=apolipoprotein A1; Apo−B=apolipoprotein B;DHA=docosahexaenoic acid; FA=fatty acids; FAw3=omega−3 fatty acids; FAw6=omega−6 fatty acids; HDL=high density lipoproteins; HDL−D=high density lipoprotein particle diameter; IDL=intermediate density lipoproteins; L=large; LA=linoleic acid; LDL=low density lipoproteins; LDL−D=low density lipoprotein particle diameter; LP=lipoprotein; M=medium; MUFA=monounsaturated fatty acids; PC1=principal component 1; PC2=principal component 2; PC3=principal component 3; PC4=principal component 4; PC5=principal component 5; PC6=principal component 6; PC7=principal component 7; PC8=principal component 8; PC9=principal component 9; PC10=principal component 10; PC11=principal component 11; PUFA=polyunsaturated fatty acids; S=small; SFA=saturated fatty acids; T2D=type 2 diabetes; VLDL=very low density lipoproteins; VLDL−D=very low density lipoprotein particle diameter; XL=very large; XS=very small; XXL=extremely large

### Webfigure 8. Characterisation of the first 11 metabolic biomarker principal components

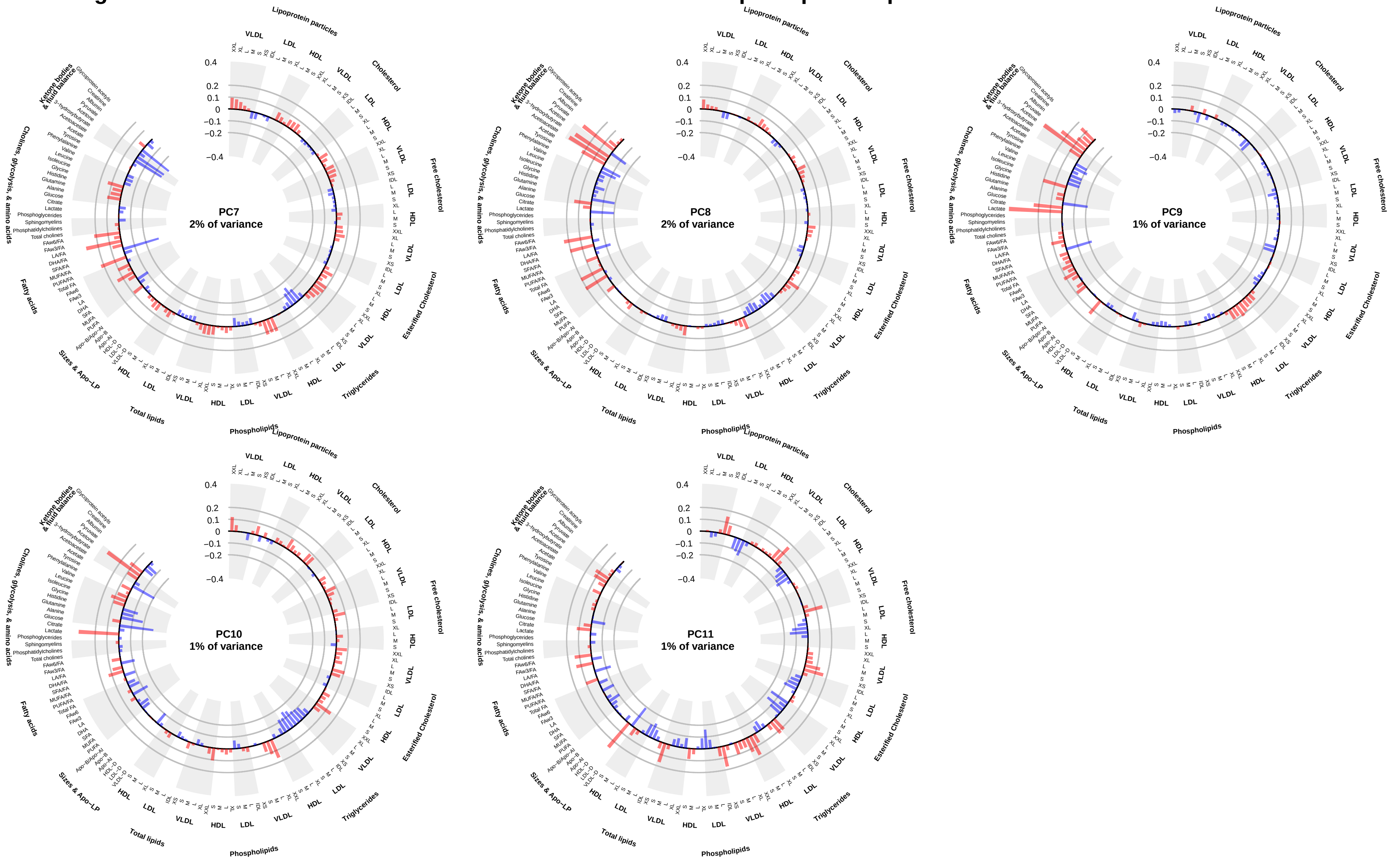


Factor loadings for each principal component are presented, representing the coefficients of the metabolic biomarkers from which each principal component was calculated. The first 11 principal components explain 90 % of the variance.
Apo−A1=apolipoprotein A1; Apo−B=apolipoprotein B; DHA=docosahexaenoic acid; FA=fatty acids; FAw3=omega−3 fatty acids; FAw6=omega−6 fatty acids; HDL=high density lipoproteins; HDL−D=high density lipoprotein particle diameter; IDL=intermediate density lipoproteins; L=large; LA=linoleic acid; LDL=low density lipoproteins; LDL−D=low density lipoprotein particle diameter; LP=lipoprotein; M=medium; MUFA=monounsaturated fatty acids; PC1=principal component 1; PC2=principal component 2; PC3=principal component 3; PC4=principal component 4; PC5=principal component 5; PC6=principal component 6; PC7=principal component 7; PC8=principal component 8; PC9=principal component 9; PC10=principal component 10; PC11=principal component 11; PUFA=polyunsaturated fatty acids; S=small; SFA=saturated fatty acids; T2D=type 2 diabetes; VLDL=very low density lipoproteins; VLDL−D=very low density lipoprotein particle diameter; XL=very large; XS=very small; XXL=extremely large

### Webfigure 9. Comparison of biomarkers measured by NMR and routine clinical chemistry assays

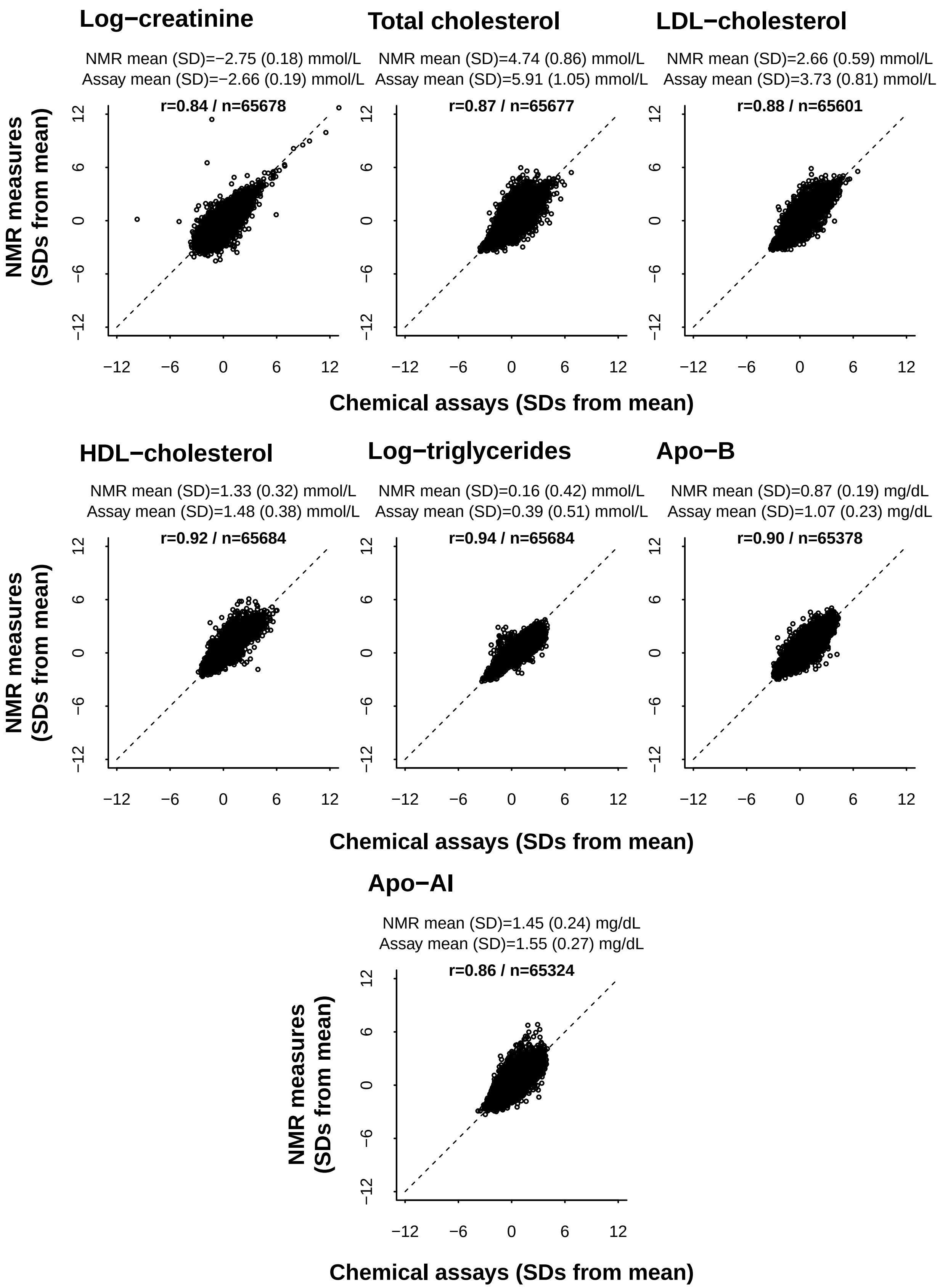


Exclusions applied as in Webfigure 1
r = Pearson's correlation coefficient between NMR measures and chemical assays.
Clinical chemistry assays were conducted on serum samples. NMR profiling was conducted on EDTA plasma samples from different aliquots.
For the cholesterol chemical assay, LDL−cholesterol was directly measured using a technique that first solubilises and extracts CM−, VLDL− and HDL−carried cholesterol, and then quantifies the remaining cholesterol carried by both IDL and LDL particles. For NMR measures, LDL−cholesterol corresponds to the sum of all the NMR−derived measures of both esterified and free cholesterol carried by both IDL and LDL particles.
Apo= apolipoprotein; NMR= nuclear magnetic resonance
